# Studying autism using untargeted metabolomics in newborn screening samples

**DOI:** 10.1101/2020.04.17.20069153

**Authors:** Julie Courraud, Madeleine Ernst, Susan Svane Laursen, David M. Hougaard, Arieh S. Cohen

**Affiliations:** Section for Clinical Mass Spectrometry, Danish Center for Neonatal Screening, Department of Congenital Disorders, Statens Serum Institut Copenhagen, Artillerivej 5, 2300 Copenhagen S, Denmark; iPSYCH, The Lundbeck Foundation Initiative for Integrative Psychiatric Research, Fuglesangs Allé 26, 8210 Aarhus, Denmark

**Keywords:** autism spectrum disorder, dried blood spots, untargeted metabolomics, newborn screening, biomarkers

## Abstract

Main risk factors of autism spectrum disorder (ASD) include both genetic and non-genetic factors, especially prenatal and perinatal events. Newborn screening dried blood spot (DBS) samples have great potential for the study of early biochemical markers of disease. To study DBS strengths and limitations in the context of ASD research, we analyzed the metabolomic profiles of newborns later diagnosed with ASD. We performed LC-MS/MS-based untargeted metabolomics on DBS from 37 case-control pairs randomly selected from the iPsych sample. After preprocessing using MZmine 2.41.2, metabolites were putatively annotated using mzCloud, GNPS feature-based molecular networking and MolNetEnhancer. 4360 mass spectral features were detected, of which 150 could be putatively annotated at a high confidence level. Chemical structure information at a broad level could be retrieved for 1009 metabolites, covering 31 chemical classes. Although no clear distinction between cases and controls was revealed, our method covered many metabolites previously associated with ASD, suggesting that biochemical markers of ASD are present at birth and may be monitored during newborn screening. Additionally, we observed that gestational age, age at sampling and month of birth influence the metabolomic profiles of newborn DBS, which informs us on the important confounders to address in future studies.

## 1. Introduction

It is estimated that autism spectrum disorder (ASD) affects more than 1% of all children worldwide [1]. ASD encompasses several neurodevelopmental disorders including autism, Asperger syndrome, pervasive developmental disorders and childhood disintegrative disorder. The etiopathology of ASD is still unclear and today ASD is diagnosed based on behavioral signs and assessment of communication skills [2,3]. How the condition should be classified is debated [4,5], as well as which tests offer the most reliable conclusions [2]. In Europe, detection services based on behavioral signs are usually accessed on average at 18 months of age, and diagnosis occurs on average at 36 months of age [6]. In this setting, early intervention is a challenge and has been reported to start in Europe at 42 months of age on average [6]. Whether behavioral impairments are reflected in the blood as biochemical abnormalities is still unsure, but the quest for biomarkers is legitimate, as they would represent a useful tool to help in the diagnosis and treatment of ASD and in understanding its underlying molecular mechanisms [7].

The main risk factors for ASD include genetic [8,9] and non-genetic factors, especially exposure during fetal life [2,10–12]. Prenatal stress could influence fetal brain development and interact with genetic predispositions thereby enhancing the risk of future psychiatric disorders [13,14]. Among prenatal outcomes, maternal infection accompanied by fever during the second trimester of pregnancy has been found to increase the risk of ASD twofold approximately [15]. Among perinatal outcomes, preterm birth (<37 weeks) and low birthweight (small for gestational age) have been associated with an increased risk of ASD as well as high frequency ventilation and intracranial hemorrhage [16]. Low Apgar scores, a score used to summarizing vital signs and assess health in newborns [17], also have recently been associated with an increased risk of developing ASD [18].

Gastrointestinal tract disorders are often reported in ASD children, along with certain foods or diets impacting the severity of symptoms [19–22]. There is a growing evidence of strong interactions between gut and brain through microbiota [23,24], and these observations support the notion that ASD is associated with metabolic malfunction such as decrease in sulphation capacity [19], and potentially connected to gut microbial populations and functions [21]. It has also been shown that many small molecules differing between normally-developing and ASD individuals likely result from microbial metabolism [21,25,26]. Recently, plasma and stool metabolites have been associated with poor communication scoring at age 3, and with good prediction of ASD by age 8 [27]. Sharon and collaborators [26] have shown that microbiome and metabolome profiles of mice harboring human microbiota predict that specific bacterial taxa and their metabolites modulate ASD behaviors. They found that taurine and 5-amiovaleric acid (5AV) had significantly lower levels in ASD mice and could show that when feeding BTBR mice either taurine or 5AV, ASD-like symptoms such as repetitive behavior and decreased social interaction could be decreased. In an intervention study, treatment with *Lactobacillus reuteri* has been shown to have beneficial effects on ASD-related social disturbances in mice [28]. In humans, intestinal microbiota transplantation has shown very promising results, both against gastrointestinal tract symptoms and ASD symptoms, granting the therapy a ‘fast-track’ status by the FDA [29]. Among the plasma metabolites showing average to good classification capacity between the treated children and the controls, sarcosine, tyramine O-sulfate and inosine 5’-monophosphate were selected as most discriminant [30]. Many of these studies postulate that microbiota-derived molecules are transported across the blood-brain-barrier, acting as neuroactive metabolites [23]. An impaired intestinal permeability or ‘leaky gut’ could also play a role in the effect of microbiota activity on psychiatric disorders [31,32]. If gut microbial metabolites of potential impact are indeed detectable in blood, this opens the door to blood-based investigations to further study and understand the metabolomic differences between ASD and non-ASD individuals in the context of gut-brain interactions.

Several studies have reported an altered metabolome associated with ASD during childhood, either in blood [33–36,26,37–46], urine [19,39,47–58] or other matrices [26,59]. However, although some biochemical markers or set of markers seem promising [7], none has yet been proven robust enough for clinical practice. Furthermore, it remains unclear at what point in life biochemical abnormalities of ASD are detectable.

To study the early role of genetic, prenatal and perinatal variables on disease development, samples need to be collected shortly after birth. However, it is not practically and ethically straightforward to draw blood from newborns prospectively. In many countries, the newborn screening programs are conducted on dried blood spots (DBS) collected a few days after birth. In Denmark, such DBS are stored in the Danish National Biobank and are available for research purposes for the last 40 years, thereby covering approximately half of the country’s population [60]. This allows researchers to alleviate the biases inherent to recruitment in prospective clinical studies and instead retrospectively retrieve the samples that are connected to the relevant metadata stored in centralized health registries.

Taking advantage of this unique resource, we here aimed at studying the strengths and limitations of DBS samples in studying early biochemical abnormalities related to ASD development using an untargeted metabolomics protocol. We compared the metabolomic profiles of newborns that have been diagnosed with ASD by age 7 (cases) to newborns that have not (controls) and investigated potential main confounders. Although no clear case-control distinction was revealed, 18 compounds repeatedly reported in the ASD literature could be detected and three mass spectral features were differentially abundant in cases and controls before FDR correction. Additionally, we observed that gestational age, age at sampling and month of birth influence the chemical profiles of neonates.

## 2. Results

### Subjects

Subjects’ characteristics are presented in Table 1 **Table**(details in Table S1).

**Table 1.** Subjects characteristics.

|  | n = 68 |  |
| --- | --- | --- |
|  | cases | controls |
| Age at 1<sup>st</sup> Jan. 2006 (median [range]) | 7.3 mo. [0.8-11.6] | 7.5 mo. [0.8-11.6] |
| Gender (girls / boys) | 7 / 25 | 8 / 28 |
| Classification of cases (ICD10)<sup>1</sup> |  |  |
| - F84.0 Childhood autism | 15 | - |
| - F84.1 Atypical autism | 6 | - |
| - F84.5 Asperger syndrome | 3 | - |
| - F84.8 Other pervasive developmental disorders | 4 | - |
| - F84.9 Unspecified pervasive developmental disorders | 10 | - |
| Gestational age (median [range]) | 40 weeks [33-41] | 39 weeks [30-42] |
| Birthweight (median [range]) | 3498 g [2210-4880] | 3490 g [977-4850] |
| Age at sampling (median [range]) | 6 days [3-9] | 6 days [4-10] |
| Age of mother at birth (median [range]) | 32.3 years [20.8-41.5] | 31.8 years [18.3-41.2] |

<sup>1</sup> ICD10 classification [3]
More details are provided in Table S1.

Cases and controls were similar in terms of gestational age (GA), birthweight, age at sampling and age of their mother at birth. The most prevalent ASD subtype was childhood autism. Most cases had only one diagnosis, but six had both unspecified pervasive development disorder and autism (either childhood autism or atypical autism). None had more than two diagnoses. Median age at first diagnosis was 5.6 years (range 1.1-7.8). Most subjects were born at term (GA ≥ 38 weeks). Only three cases and two controls were born preterm.

### Molecular Network analysis

From all features for which a MS2 spectrum has been acquired (2217 features over 4360) a feature-based molecular network was computed via GNPS. Annotation could be retrieved for 150 features (3.4%) of which 103 by matching to GNPS libraries (annotation level 2), and 47 by matching to our in-house library using Trace Finder (annotation level 1, Table S2). Using the MolNetEnhancer workflow [61], putative chemical structural information at the chemical class level, corresponding to a level 3 annotation, could be retrieved for an additional 859 features. Hence, nearly 46% (1009) of the mass spectral features could be putatively annotated at a level 1 to 3 (Table S2). Annotation covered 31 chemical classes including 53 subclasses and 116 direct parents, such as medium-chain fatty acids, phosphatidylcholines, nucleotides, amino acids, bile acids, steroids, acyl carnitines and catecholamines.

Molecular families (independent clusters of nodes) from the 15 predominant putatively annotated chemical classes are presented in Figure 1 (see details in Table S2). Plotting the average intensities in the three groups (cases, controls, paper blanks) as well as fold change values (or p-values) on the network nodes allowed for a quick overview of the molecular families with potential biological relevance (See the example of bile acids in Figure 2). This analysis showed the potential of DBS in covering various chemical classes and the power of feature-based molecular network analyses and related metabolome mining tools in expanding the interpretability of complex untargeted metabolomics data.

**Figure 1.**
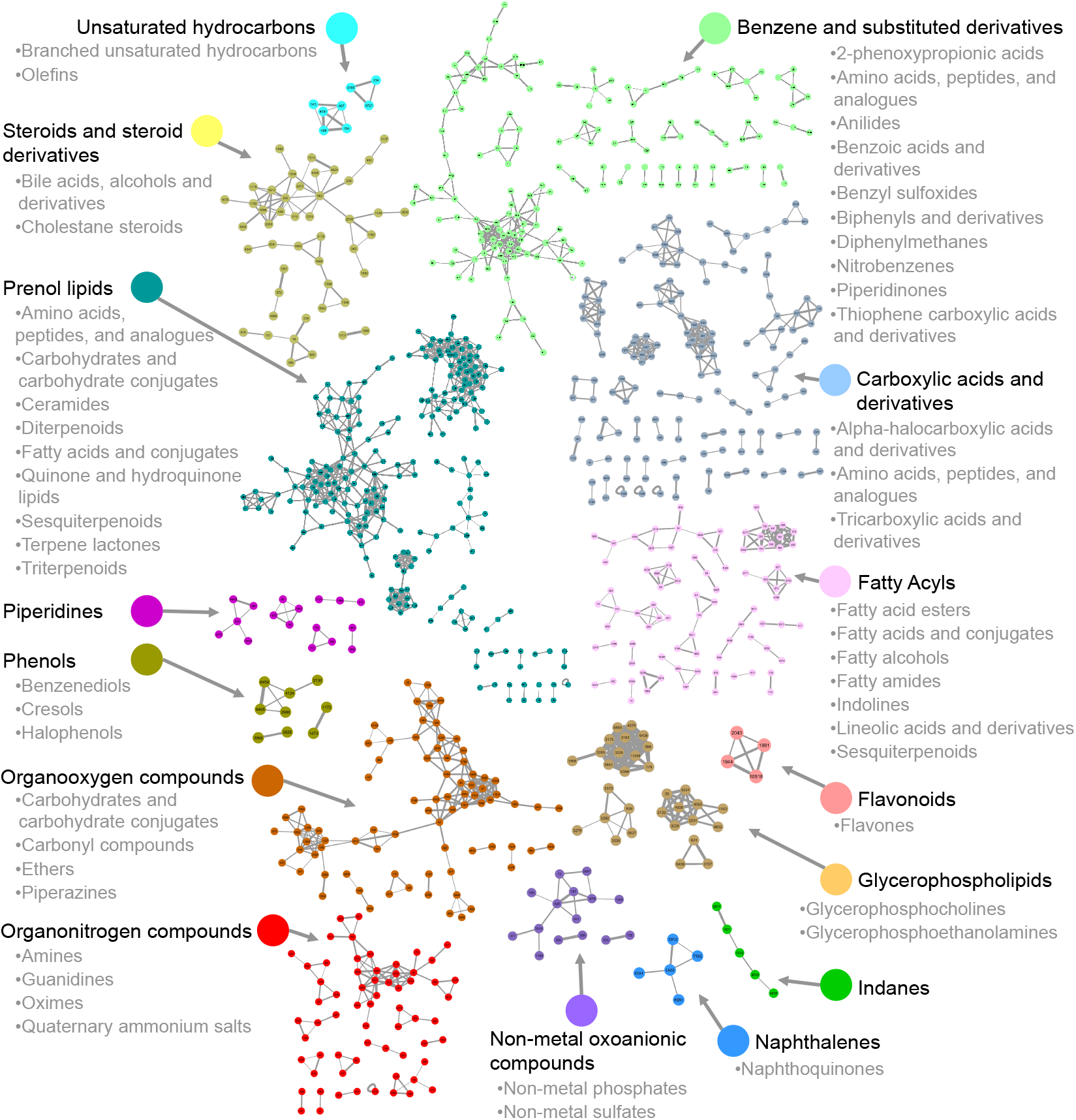
Feature-based molecular network displaying the 15 predominant putative chemical classes and their subclasses. Nodes represent mass spectral features and are used as a proxy for a metabolite. Connected nodes represent high tandem mass spectral similarity, and thus high chemical structural similarity. The thickness of the grey edges connecting nodes varies according to the cosine score representing to what extent two connected metabolites are chemically similar (based on MS2 spectra, from 0.7: less similar and thin edge to 1.0: identical and thick edge). The name of annotated metabolites (levels 1 and 2), details on chemical classes with fewer than 4 metabolites (absent on this figure), chemical classification scores [61], all unknowns, and group intensities for all features (average, standard deviations) are detailed in Table S2. Note that data represent a summary of most predominant classes per molecular family retrieved through either GNPS spectral library matching or *in silico* structure annotation and may contain false positives.

**Figure 2.**
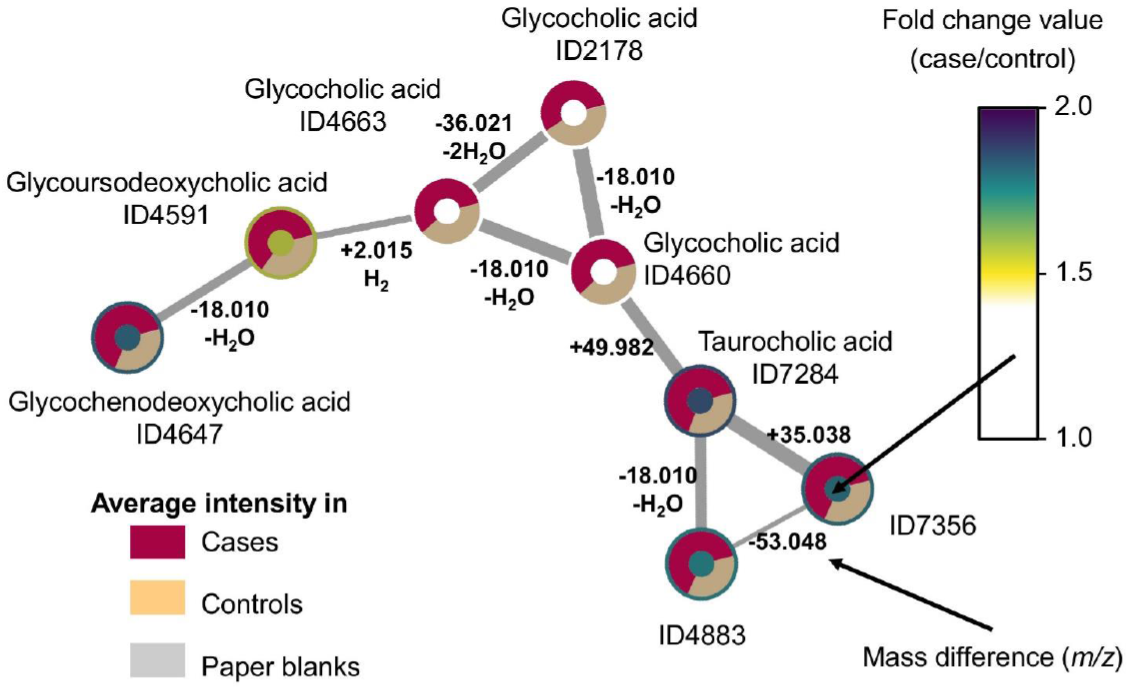
Network of molecular features putatively annotated as bile acids with average group intensities, fold change values, mass differences and cosine scores displayed. Molecular family #75 is composed of eight bile acid structural analogues (see details in Table S2). Coloring according to the fold change values makes it easier to spot the families with differential abundance in cases vs. controls. Displaying average intensities for the three groups (cases, controls, paper blanks) allows for a quick control of the matrix signals (paper blanks, here none of the features were detected in the matrix) and confirmation of fold change. On edges, while the thickness of the connection represents to what extent two metabolites are chemically similar, the mass difference is essential to support annotation as it translates into how molecules differ from one another (e.g. water loss, conjugation, adducts, etc.).

### Statistical analyses

Principal component analysis revealed that repeated pool injections clustered satisfactorily showing that the LC-MS/MS data acquisition was of acceptable quality (Figure 3). When looking at the two groups (cases/controls), no clear separation was observed, even after removal of outliers (Figure 3).

**Figure 3.**
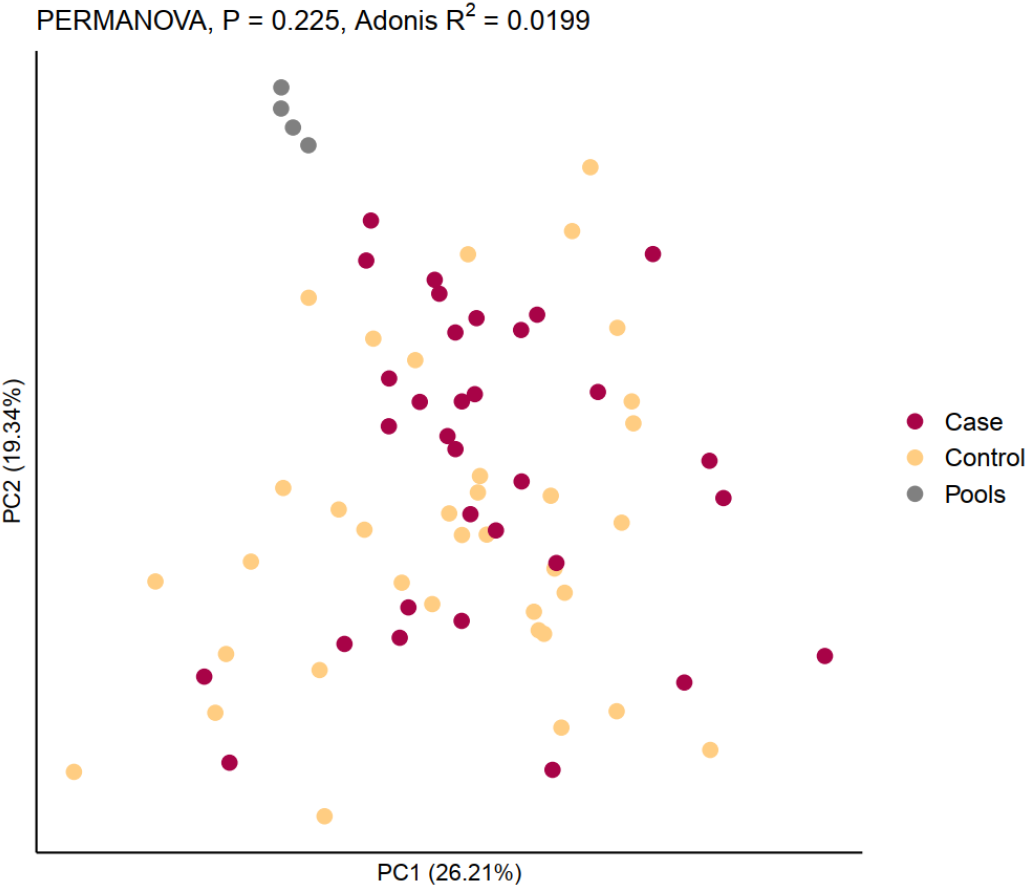
Principal component analysis of the 68 samples after outlier removal reflecting the distribution of cases and controls. Each sphere represents one sample. Axes are principal components 1 (x) and 2 (y) explaining 26.21% and 19.34% of the variation in the data, respectively. The four replicated pool injections cluster satisfactorily. Coloring reflects the type of samples, i.e. cases, controls and four replicated pool injections. No clear distinction between cases and controls can be observed (PERMANOVA Adonis R^2^ = 0.0199, P-value = 0.226).

PERMANOVA (Figure 3 and 4, Table S3) revealed that the variance in the data was not significantly explained by the grouping (cases/controls) (Adonis R^2^ = 0.0199, P-value = 0.226), even when distinguishing subtypes of ASD (Adonis R^2^ = 0.123, P-value = 0.546, see Table 1 for details on subtypes of ASD). Similarly, the gender and birthweight did not significantly explain the variance in the data (Adonis R^2^ < 0.02, P-value > 0.05). However, variation in the data explained by gestational age (Adonis R^2^ = 0.0429, P-value = 0.021), age at sampling (Adonis R^2^ = 0.0425, P-value = 0.016) and especially month of birth (Adonis R^2^ = 0.272, P-value = 0.001) was significant (Figure 4).

**Figure 4.**
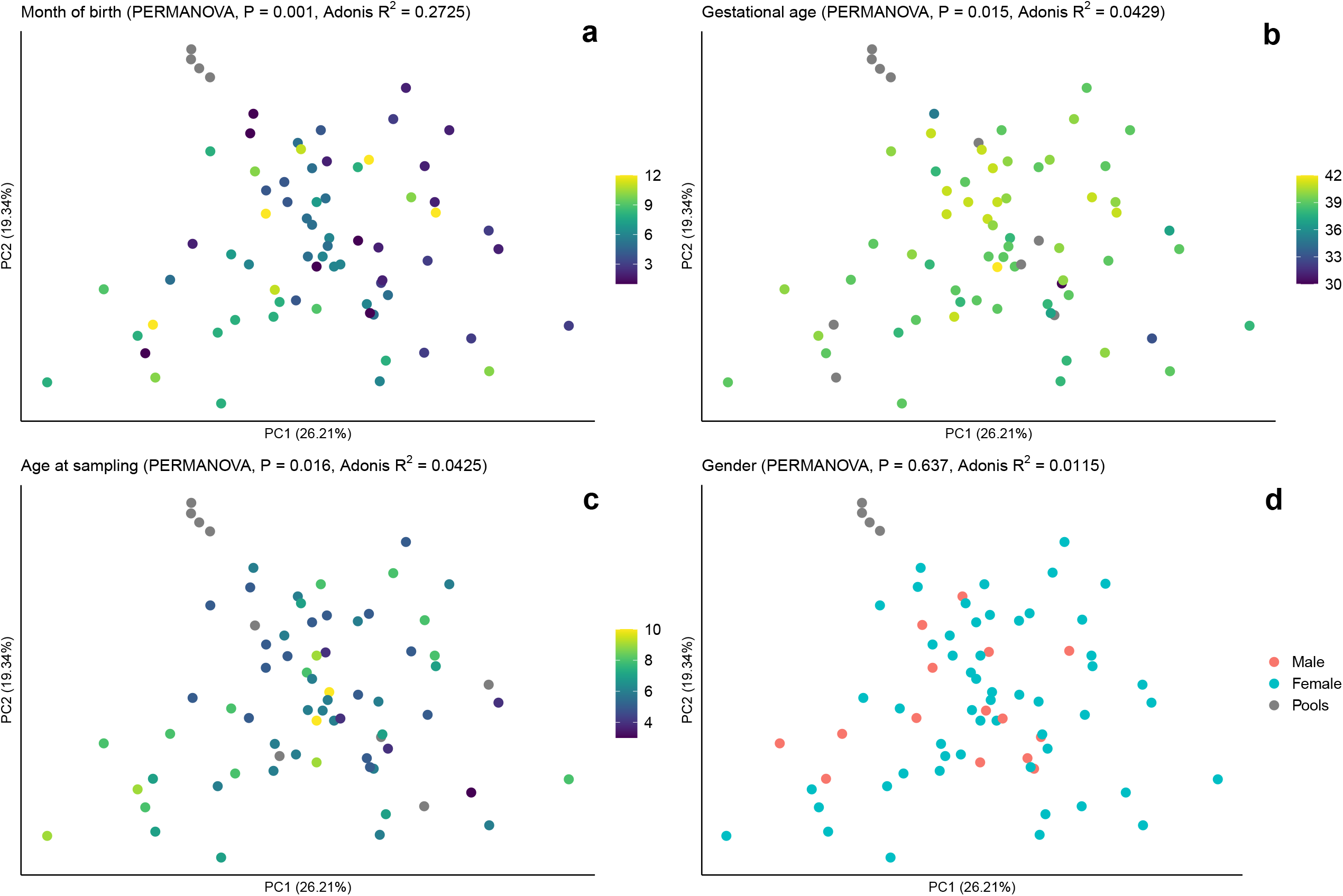
Principal component analysis of the 68 samples after outlier removal. Each sphere represents one sample. Axes are principal components 1 (x) and 2 (y) explaining 26.21% and 19.34% of the variation in the data, respectively. (a) Coloring reflects the month of birth. The largest variation in the data (27%) could be explained by the month of birth (Adonis R^2^ = 0.2725, P-value = 0.001), with children born during the winter months being more similar to each other than children born during the summer months, (b) Coloring reflects gestational age. Over 4% of the variation in the data could be explained by gestational age (Adonis R^2^ = 0.0429, P-value = 0.015), (c) Coloring reflects age at sampling. Over 4% of the variation in the data could b explained by age at sampling (Adonis R^2^ = 0.0425, P-value = 0.016), (d) Coloring reflects gender. No significant effect on the metabolome by gender was observed (Adonis R^2^ = 0.0115, P-value = 0.582).

Results of univariate analyses and fold change analysis were carefully scrutinized feature by feature. Considering our small sample size and potential pitfalls inherent to untargeted metabolomics related to contaminants or integration errors, we thought essential to inspect each result to eliminate false positives and spurious findings. Our inspection consisted of a five-step logic starting with peak integration and shape quality (MZmine). We then plotted all individual intensity values to assess whether the case/control difference was driven by four or fewer samples. If not, we reported the extent of missing values in each group, checked the consistency of replicated pool injections, and finally checked whether the feature was present in the feature-based molecular network, annotated as a contaminant or in a node cluster with such annotation (Table S4). A large proportion of the inspected features were excluded based on these criteria, showing the importance of such a verification in order not to pursue spurious findings in future studies.

Among the 24 features with a fold change (case/control) value <0.5 or >2.0, only one passed manual inspection (Table 2, the full table is in Table S4).

**Table 2.**
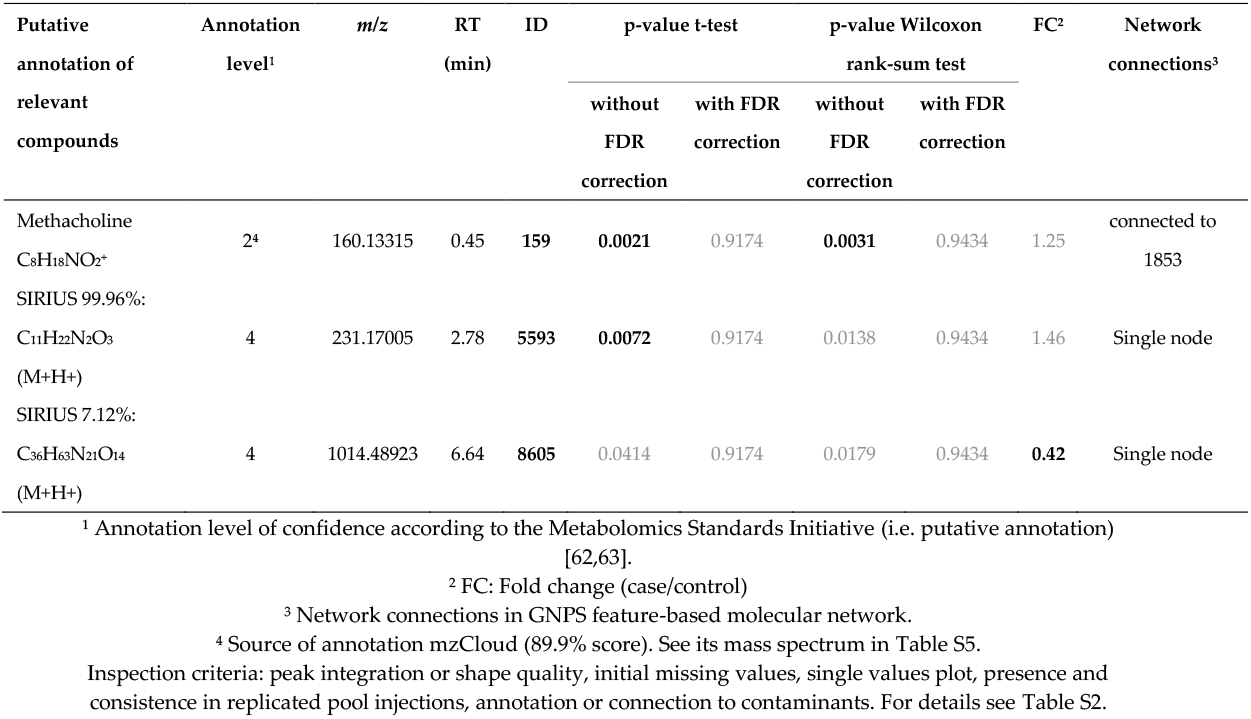
Differentially abundant features in univariate analyses without FDR correction (p<0.01, two features) and/or with high fold change (one feature) meeting inspection criteria

Eluting quite late (RT = 6.64 min, ID8605), this relatively hydrophobic compound had a detected *m/z* of 1014.4892 and was not connected to any other node in the network analysis (see its mass spectrum in Table S5). It could not be annotated, but the algorithm of SIRIUS+CSI:FingerID pointed at a raw formula of C_36_H_63_N_21_O_14_ ([M+H]+, only 7.12% scoring). This compound was more than twice as intense in controls as in cases (FC 0.42, average intensity in cases 2.73E+05 and controls 7.51E+05), and would need further investigation, especially as it was not detected in many samples (Table S4). A MASST search was performed, however the feature with *m/z* 1014.4892 was not found in any of the public datasets on GNPS.

No feature was significantly differentially abundant in cases and controls according to the univariate analyses with FDR correction for multiple comparisons (p-values in Table 2).

Features that were differentially abundant before FDR correction are presented in Table 2. As a high proportion of features were deemed irrelevant after inspection, we are presenting only the two relevant features that passed our quality-control criteria. The full list and inspection details can be found in Table S4. Methacholine was found to be significantly more abundant in cases when compared to controls (average intensity in cases 4.41E+07 and controls 3.94E+07) both when using a t-test (p = 0.0021) and a Wilcoxon rank-sum test (p = 0.0031). The corresponding node (ID159) in the network analysis was connected to another node with a mass difference of −0.036 *m/z* (225 ppm) which could not be annotated. None of the applied metabolome mining tools was able to retrieve chemical structural information for the second compound significantly more abundant in cases than in controls (ID5593, *m/z* 1014.4892, average intensity in cases 5.71E+05 and controls 4.35E+05). SIRIUS+CSI:Finger ID predicted a molecular formula of C_11_H_22_N_2_O_3_ (M+H+, 99.96% scoring). Its RT of 2.78 min could indicate a medium polarity with a logP between −1.0 and 0.5 when compared to tryptophan (RT 2.56 min, HMDB experimental logP −1.06) and hippuric acid (RT 3.04 min, HMDB experimental logP 0.31).

Among the 273 compounds reported in two recent reviews [7,64], 22 were cited at least three times, of which 18 could be linked to features in our study after manual verification (Table 3, Table S6).

**Table 3.**
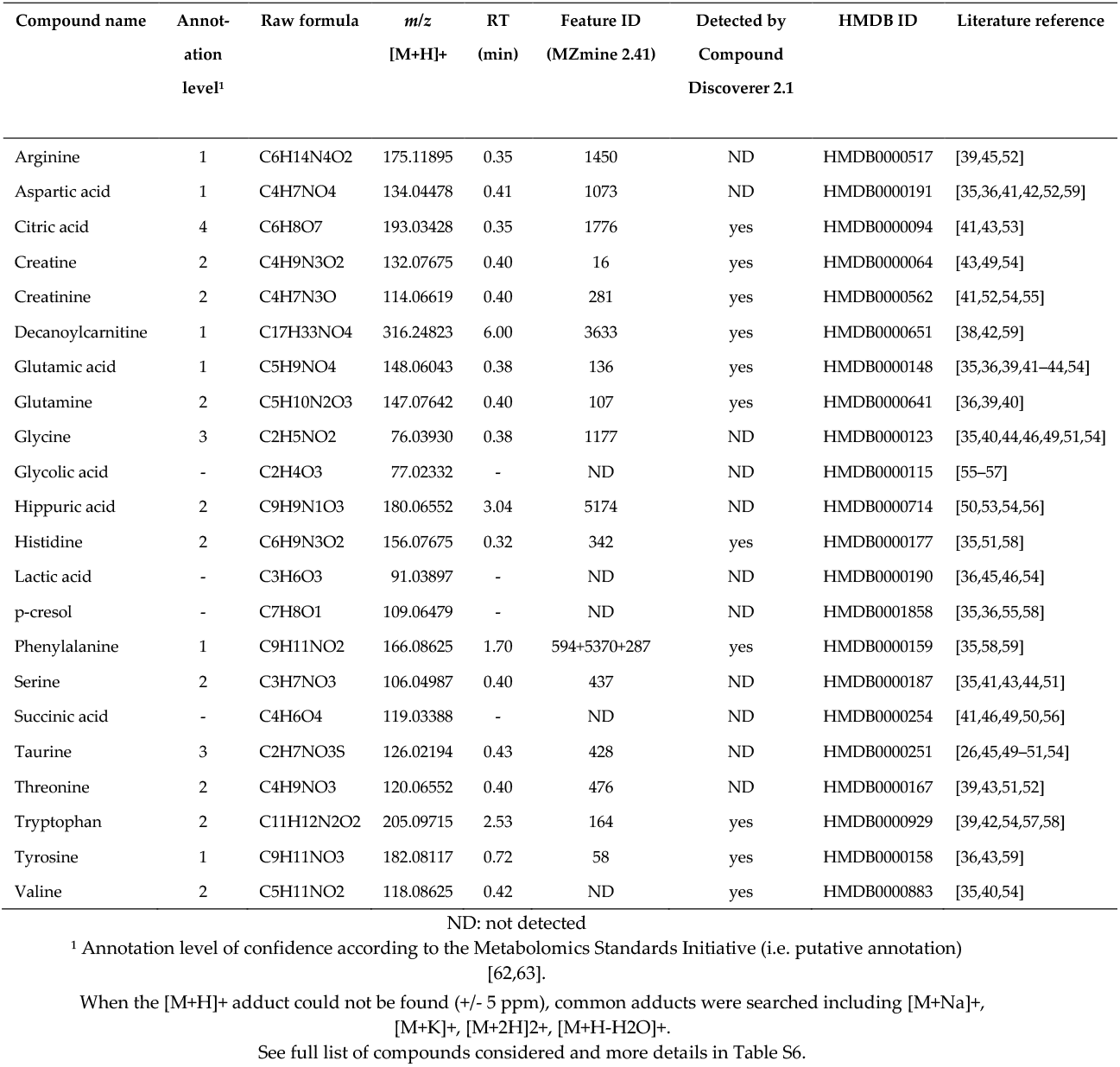
Compounds reported in the literature three or more times as being associated with ASD.

| Compound name | Annot-<br>ation<br>level<sup>1</sup> | Raw formula | $m/z$<br>[M+H]<sup>+</sup> | RT<br>(min) | Feature ID<br>(MZmine 2.41) | Detected by<br>Compound<br>Discoverer 2.1 | HMDB ID | Literature reference |
| --- | --- | --- | --- | --- | --- | --- | --- | --- |
| Arginine | 1 | C<sub>6</sub>H<sub>14</sub>N<sub>4</sub>O<sub>2</sub> | 175.11895 | 0.35 | 1450 | ND | HMDB0000517 | [39,45,52] |
| Aspartic acid | 1 | C<sub>4</sub>H<sub>7</sub>NO<sub>4</sub> | 134.04478 | 0.41 | 1073 | ND | HMDB0000191 | [35,36,41,42,52,59] |
| Citric acid | 4 | C<sub>6</sub>H<sub>8</sub>O<sub>7</sub> | 193.03428 | 0.35 | 1776 | yes | HMDB0000094 | [41,43,53] |
| Creatine | 2 | C<sub>4</sub>H<sub>9</sub>N<sub>3</sub>O<sub>2</sub> | 132.07675 | 0.40 | 16 | yes | HMDB0000064 | [43,49,54] |
| Creatinine | 2 | C<sub>4</sub>H<sub>7</sub>N<sub>3</sub>O | 114.06619 | 0.40 | 281 | yes | HMDB0000562 | [41,52,54,55] |
| Decanoylcarnitine | 1 | C<sub>17</sub>H<sub>33</sub>NO<sub>4</sub> | 316.24823 | 6.00 | 3633 | yes | HMDB0000651 | [38,42,59] |
| Glutamic acid | 1 | C<sub>5</sub>H<sub>9</sub>NO<sub>4</sub> | 148.06043 | 0.38 | 136 | yes | HMDB0000148 | [35,36,39,41–44,54] |
| Glutamine | 2 | C<sub>5</sub>H<sub>10</sub>N<sub>2</sub>O<sub>3</sub> | 147.07642 | 0.40 | 107 | yes | HMDB0000641 | [36,39,40] |
| Glycine | 3 | C<sub>2</sub>H<sub>5</sub>NO<sub>2</sub> | 76.03930 | 0.38 | 1177 | ND | HMDB0000123 | [35,40,44,46,49,51,54] |
| Glycolic acid | - | C<sub>2</sub>H<sub>4</sub>O<sub>3</sub> | 77.02332 | - | ND | ND | HMDB0000115 | [55–57] |
| Hippuric acid | 2 | C<sub>9</sub>H<sub>9</sub>N<sub>1</sub>O<sub>3</sub> | 180.06552 | 3.04 | 5174 | ND | HMDB0000714 | [50,53,54,56] |
| Histidine | 2 | C<sub>6</sub>H<sub>9</sub>N<sub>3</sub>O<sub>2</sub> | 156.07675 | 0.32 | 342 | yes | HMDB0000177 | [35,51,58] |
| Lactic acid | - | C<sub>3</sub>H<sub>6</sub>O<sub>3</sub> | 91.03897 | - | ND | ND | HMDB0000190 | [36,45,46,54] |
| p-cresol | - | C<sub>7</sub>H<sub>8</sub>O<sub>1</sub> | 109.06479 | - | ND | ND | HMDB0001858 | [35,36,55,58] |
| Phenylalanine | 1 | C<sub>9</sub>H<sub>11</sub>NO<sub>2</sub> | 166.08625 | 1.70 | 594+5370+287 | yes | HMDB0000159 | [35,58,59] |
| Serine | 2 | C<sub>3</sub>H<sub>7</sub>NO<sub>3</sub> | 106.04987 | 0.40 | 437 | ND | HMDB0000187 | [35,41,43,44,51] |
| Succinic acid | - | C<sub>4</sub>H<sub>6</sub>O<sub>4</sub> | 119.03388 | - | ND | ND | HMDB0000254 | [41,46,49,50,56] |
| Taurine | 3 | C<sub>2</sub>H<sub>7</sub>NO<sub>3</sub>S | 126.02194 | 0.43 | 428 | ND | HMDB0000251 | [26,45,49–51,54] |
| Threonine | 2 | C<sub>4</sub>H<sub>9</sub>NO<sub>3</sub> | 120.06552 | 0.40 | 476 | ND | HMDB0000167 | [39,43,51,52] |
| Tryptophan | 2 | C<sub>11</sub>H<sub>12</sub>N<sub>2</sub>O<sub>2</sub> | 205.09715 | 2.53 | 164 | yes | HMDB0000929 | [39,42,54,57,58] |
| Tyrosine | 1 | C<sub>9</sub>H<sub>11</sub>NO<sub>3</sub> | 182.08117 | 0.72 | 58 | yes | HMDB0000158 | [36,43,59] |
| Valine | 2 | C<sub>5</sub>H<sub>11</sub>NO<sub>2</sub> | 118.08625 | 0.42 | ND | yes | HMDB0000883 | [35,40,54] |

ND: not detected
<sup>1</sup> Annotation level of confidence according to the Metabolomics Standards Initiative (i.e. putative annotation) [62,63].
When the [M+H]<sup>+</sup> adduct could not be found ( ± 5 ppm), common adducts were searched including [M+Na]<sup>+</sup>, [M+K]<sup>+</sup>, [M+2H]<sup>2+</sup>, [M+H-H<sub>2</sub>O]<sup>+</sup>.
See full list of compounds considered and more details in Table S6.

## 3. Discussion

To assess the potential of newborn DBS to study early biochemical markers of ASD shortly after birth, we compared DBS samples from newborns that have later on been diagnosed with ASD to newborns that have not. Our study showed the capacity of untargeted metabolomics as an analytical tool applied to biobanked DBS samples to cover several metabolites relevant to ASD, thus suggesting that biochemical markers of ASD are present at birth and could be targeted during neonatal screening. In addition, our method pinpointed other factors which have a strong influence on the metabolic profile of newborn DBS, such as gestational age, age at sampling and month of birth, and which are important to consider when designing metabolomic studies in neonatal, biobanked DBS.

One study from 2013 was performed on newborn DBS samples from 16 autistic children and assessed 90 biomarkers (not only small molecules) using immunoassays [65] of which three sets of five were associated with ASD. Another study was performed on DBS but in ASD children (n=83, age 2-10 years) and was targeting 45 metabolites [38], of which 9 were significantly higher in ASD children. However, the potential of DBS in untargeted metabolomics studies has not yet been fully studied, and never in the context of ASD (see recent reviews [7,64]).

Among the 22 compounds that had been repeatedly (≥3 times) reported in the literature to be involved in ASD, 18 could be putatively annotated in our study, showing that our analytical pipeline covers many relevant metabolites, including some specific to gut microbiota activity. Despite thorough curation and inspection of the acquired data, no feature was significantly differentially abundant in cases and controls after FDR correction. This shows that a bigger sample size will be required for the study of ASD using newborn DBS along with appropriate consideration of the confounders specific to these samples to reduce their impact.

Among the hits and interesting findings of our study, we could show that a methacholine structural analogue could be a relevant marker for ASD, as it was found at a higher -although not significant-abundance in newborns that have been diagnosed with ASD by age 7. Methacholine is a choline ester drug acting as non-selective muscarinic receptor agonist. It is mainly known as methacholine chloride for its use in assessing bronchial hyper-reactivity in asthmatic patients. Although muscarinic receptors were not associated with ASD in children [66], lower estimates of ASD risk among children exposed during fetal life to muscarinic receptor 2 agonists were reported [67]. Higher abundance of methacholine in DBS of ASD cases, as seen in our study, would therefore not be easily explained and demand further investigation. However, detecting a drug metabolite such as methacholine in newborn samples is unexpected, thus it is more likely that this feature is an endogenous choline ester with similar fragmentation behavior to methacholine.

Two other unknown features would benefit from being monitored in future studies. One relatively hydrophobic compound (ID8605, *m/z* 1014.4892) showed an important fold change (much lower in cases) but was not detected in many samples maybe due to low intensities. The second compound (moderately polar, ID5593, *m/z* 1014.4892, C11H22N2O3) was significantly higher in cases before FDR correction and detected in more than 65% of samples.

We have shown that gestational age, age at sampling and month of birth are strong drivers of metabolomic profiles in newborn DBS samples. This demonstrates the importance of considering these confounders when designing a future study using such samples.

Prematurity has been involved in numerous adverse health outcomes [68] and metabolic maturity has previously been shown to be reflected in the blood and other matrices of infants after birth [69,70]. Although, in the present study, only three cases and two controls were premature (<38 weeks of gestational age), we saw a significant effect of gestational age on the metabolomic profile of newborns thus showing that gestational age is an important factor to be controlled for in newborn DBS studies.

Similarly, we found that age at sampling has a significant impact on the newborn blood metabolome. From 3 to 10 days of age, only one week has passed, and yet fundamental metabolic changes occur in the newborn possibly in connection with post-natal nutrition, the maturation of the newborn’s microbiome as well as environmental conditions (healthcare, hospital vs home, etc.). The endogenous anabolism/catabolism balance is in itself a strong variable to consider at that age. From 2009 onwards, the Danish newborn screening program has indeed chosen to standardize the age at DBS sampling to 48 to 72 hours to optimize the window where potential inborn errors of metabolism would be detected best and as early as possible since quick intervention is essential in such cases [71]. The iPsych sample was based on diagnoses of psychiatric disorders recorded in Danish health registry in 2012 [72]. Such diagnoses are often given after several years of age, which is why the iPsych sample included subjects born latest in 2005, year at which the age at sampling was not so narrowly standardized.

Another major change that occurs in newborns at birth and in the following days is the gut maturation and its further colonization by microbes [73]. This topic has been under expanding attention in the last decade, and the development and involvement of gut microbiota in neurodevelopment is being scrutinized extensively [23]. The exact dynamics of the microbiota development in the placenta and during the first days of life is still uncertain [73–75], as well as to what extent its activity can be reflected in the blood. A recent study has shown that gut microbial alpha-diversity can be predicted from the human blood metabolome [76] suggesting that microbial metabolites explain a significant amount of the variation in the human blood metabolome. Thus, although sampled at an early stage in life, it is plausible that microbial metabolites mediating health may be found in dried blood spots from newborns [70]. Studying both fecal and blood samples will be essential to answer questions related to the impact of gut microbes on the gut-brain axis, especially in the context of psychiatric disorders where the brain is the main organ concerned but indeed located quite far from the gut. Microbial metabolites would necessarily need to travel in the blood (or lymph) and through the blood-brain barrier to interact with the brain. In our study, some detected metabolites could partly derive from gut microbiota activity such as DL-Indole-3-lactic acid (ID3461, [77,78]), taurine (ID428, level 3, [26]), various bile acids (Table S2, [59]), or inosine 5’-monophosphate (ID1133, level 3, [30]).

Lastly, we found that month of birth explains a significant variation in metabolomic profiles of newborns (Figure 4). Whether there is a yearly cyclic pattern or whether our findings are specific to 2005 remains to be determined. Explanations could include aspects related to pregnancy conditions varying along the year such as diet, weather conditions and sun exposure (e.g. impact on vitamin D levels, type and extent of physical and social activities, mood and stress [79]), exposure to “seasonal” infectious diseases (e.g. influenza), exposure to varying air quality (e.g. pollution or pollens [80]), as well as sample storing conditions which might fluctuate over the year (e.g. sample transport at higher temperatures during summer).

Gender and birthweight were not found to explain a significant part of the variance in the metabolomic profiles of newborn DBS samples in our study, despite the obvious connection between gestational age and birthweight. The gender misbalance which reflects the gender disparity in ASD (a quarter were girls) and small sample size could explain this finding. Some studies have indeed reported that the profile of newborn girls and boys differed in, for instance, blood amino acids and acylcarnitines [81], as well as urine profiles [82]. Despite our finding, we believe that gender and birthweight should be adjusted for and taken into consideration when designing metabolomics studies in newborns. Several of the tested confounders are inter-connected with, for instance, reports of more males being born preterm [83] and females being born lighter [84], both associations being explained by mechanisms that are likely to be reflected in the metabolome such as inflammatory response and insulin resistance, respectively.

### Limitations and strengths

To minimize the use of highly valuable and rare samples, we analyzed only 37 pairs of cases and controls in this study aiming at assessing the potential of DBS samples in ASD research. Despite the small sample size that did not confer enough statistical power for pinpointing strong marker metabolites of ASD, we could detect numerous metabolites associated with ASD in previous studies and identify a number of confounders to be considered in future untargeted metabolomics study using newborn DBS. Other confounders not evaluated in our study will need to be assessed in future studies, including metabolic changes in DBS associated with time and storage conditions. Hematocrit variation could not be measured in our study as we had access to only one punch of paper and did not have the possibility to measure a surrogate marker such as potassium in the same punch as done by others [85]. Furthermore, metabolites detected in this study are inherently reflective of sampling protocols, including extraction protocols and MS acquisition parameters and should be interpreted within these limitations.

## 4. Materials and Methods

### Materials

Methanol (MeOH), acetonitrile (ACN), isopropanol (IPA), water (H2O) and formic acid (FA) were of Optima™ LCMS-grade and were purchased from Thermo Fisher Scientific (Waltham, MA, USA). Stable-isotope-labeled internal standards (IS) from the NeoBase Non-derivatized MSMS kit (PerkinElmer, Waltham, MA, USA) were used. The exact list of compounds is provided in Table S7.

### Subjects and samples

Samples were drawn from children from the iPsych case-cohort sample [72]. Eligible cases were defined as born in 2005 and with a diagnosis of autism spectrum disorder (ICD10 F84.0, F84.1, F84.5, F84.8 and/or F84.9) [3] by the date of registry data extraction (2012). Out of these eligible cases, 37 children were randomly selected for the study. Children matching the cases’ gender and date of birth and without a diagnosis of psychiatric disorder were selected as eligible controls, of which one was randomly selected for each pair (37 controls). Other metadata such as GA, birthweight, age at sampling, month of birth, mother’s age at birth, and date of diagnosis, were also collected (when available).

Sample size was chosen for several reasons: 1) the unknown variation of metabolites in DBS made power calculations impossible, 2) batch effect is a common technical challenge in metabolomics, and analyzing all samples on one single 96-well plate was expected to reduce technical variability, 3) DBS are highly precious samples.

DBS are whole blood from newborns, aged between 48 and 72 hours, blotted onto Ahlstrom #226 filter paper and left to dry for at least 3 hours at room temperature before being sent by mail at ambient temperature to the Department of Congenital Disorders at the Statens Serum Institut in Copenhagen. Subsequent to being used in the newborn screening program the samples are biobanked in the Danish National Biobank (www.nationalbiobank.dk) at −20°C until they are retrieved for further research analysis.

### Ethics approval

The Danish ethical committee approved the project (1-10-72-287-12). All blood samples can be stored without explicit informed consent according to Danish law, and be reused for additional analysis provided that projects are approved by the Research Ethics Committee and the Danish Data Protection Agency. Although research projects require informed consent, the Research Ethics Committee can waive this requirement if samples are anonymously processed, and if the projects do not imply any health-related risk or burden to the subjects [86].

### Sample extraction

A punch of 3.2-mm diameter was collected from each DBS using a Panthera-PuncherTM 9 blood spot punching system (PerkinElmer) directly into a MicroPlate, non-coated 96-well clear polystyrene plate (PerkinElmer). 100 μL of IS in extraction buffer were added to each well. The IS were labelled amino acids (AA IS) and acylcarnitines (AC IS) diluted in 80% methanol (i.e. dilution factor of 1:330, concentrations in Table S7). The plate was heat-sealed and shaken for 45 min at 750 rpm at 25°C in a PHMP-4 incubator. Then it was centrifuged for 30 min at 4000 rpm at 4 °C.

All the transferring steps were performed on a Microlab STAR line automated liquid handling workstation using Venus software (Hamilton, Bonaduz, Switzerland).

The supernatant (75 μL) was transferred to a hard-shell 96-well polypropylene PCR plate (Bio-Rad) and dried down with nitrogen 60 L/min at 25 °C for 1 hour on an EVX-192 (Apricot Designs Evaporex). The residue was reconstituted in 75 μL 2.5% methanol, shaken for 15 min at 750 rpm at 25°C in a PHMP-4 incubator, and centrifuged 10 min at 4000 rpm at 4 °C. 65µL were transferred to a hard-shell 96-well polypropylene PCR plate (Bio-Rad), heat-sealed, and centrifuged again for 5 min at 3000 rpm at 4 °C. The method from sample preparation to MS acquisition is also available as a table according to the guidelines for standardization of LCMS method reporting [87] with adaptation to metabolomics (Table S7).

### Quality assurance

LC-MS/MS instrument performance was controlled by analyzing 4 pooled extracts, 8 solvent blanks, and 3 paper blanks at regular intervals. Pooled extracts were made of 5 µL of reconstituted extract from each of the samples (cases and controls only, total of 370 µL divided in four wells spread across the plate) and were used to assess the consistency of extraction and data acquisition. Solvent blanks were used to check for carry over and instrument noise, while paper blanks (a punch of paper extracted like a sample) were used to monitor matrix signals from the paper. Internal standards were used to control the quality of the extraction, elution, and signal acquisition. Paired cases and controls were injected after one another but in a random order (first case, then control, or vice-versa). Pairs were randomized over the plate.

### Liquid chromatography

The samples were injected using an autosampler with stack cooler (Open Autosampler UltiMate OAS-3300TXRS (Thermo Fisher Scientific)) and eluted through a Waters Acquity UPLC BEH C18 column (130 Å, 2.1 mm x 50 mm, 1.7-µm particles) preceded by a Waters Acquity UPLC BEH C18 VanGuard pre-column, 130 Å, 2.1 mm x 5 mm, 1.7-µm particles) using a Transcend II, LX-2 with UltiMate pumps (Thermo Fisher Scientific). The pressure limits were set at 0.0 – 1034.0 bar.

The mobile phase consisted of solvent A (97.31% H2O, 1.25% ACN, 1.25% MeOH and 0.2% FA), and B (2.49% H2O, 48.66% ACN, 48.66% MeOH and 0.2% FA). The Wash1 solvent was mobile phase A and the Wash2 solvent mix was 25:25:25:25 v/v MeOH:IPA:H20:ACN + 0.2% FA. The gradient (0.25 mL/min) started with 100% A : 0% B. After 0.5 min, we applied a gradient ramp to 0% A: 100% B over 8.5 minutes followed by a 0.5-min flow ramp up to 0.9 mL/min and 5 minutes of 100% B. At 15 min, the column was equilibrated for 5.5 min with 100% A. At 17.5 min, the flow was changed back to 0.25 mL/min over 0.5 min. The total run time was 20.5 minutes, including 10 min sample run time and 10.5 min column wash and equilibration. The column temperature was maintained at 60.0°C using a hot pocket column heater and the samples in the autosampler were kept at 4°C throughout the analysis. The data was acquired in profile mode from 0.20 min and over 9.80 min.

### Mass spectrometry

All samples were injected once and analysed in data dependent acquisition mode. The Q-Exactive Orbitrap mass spectrometer (Thermo Fisher Scientific) was operated with a heated electrospray ionization source (HESI) in positive mode. The instruments were controlled using TraceFinder 4.1 Clinical Research and Aria MX (Thermo Fisher Scientific). Mass range in MS full scan mode was set to 70 to 1050 *m/z* with a resolution of 35,000. Automatic gain control was set to 1.106, and maximum injection time at 100 ms. For data dependent-MS2 the resolution was set to 17,500. Automatic gain control was set to 1.105, and maximum injection time at 50 ms. Loop count was 5, isolation window 1.5 *m/z* and the stepped NCE 17.5, 35 and 52.5 eV. The spectrum data type was set to Profile. In data dependent settings the Apex trigger was set to 2 to 7 s with 15s dynamic exclusion and charge exclusion on 3-8 and >8. Diisooctylphtalate (391.28429 *m/z*) was selected as lock mass. Other settings included the sheath gas pressure (N2, 32 psi), the auxiliary gas flow and temperature (N2, 8 arb. units, 350°C), the S-lens radio frequency level (50.0%), the ion source temperature (350°C), and the spray voltage (3.8 kV between 0-9.8 min and 1.0 kV between 9.8-10 min).

### LC-MS/MS data preprocessing

After conversion to .mzML (centroid) using MSConvertGUI v3.0 (ProteoWizard Software Foundation, Palo Alto, CA, USA) [88], raw files were pre-processed using MZmine v2.41.2 [89,90]. All setting details are provided in the batch .xml file (Document S1). Briefly, data were cropped based on retention time (RT) 0.27-9.80 min. Masses were detected with a noise threshold of 10,000 for MS1 and of 0 for MS2. The chromatogram was built using the ADAP module [91], with minimum 7 scans per peak, a group intensity threshold of 10,000, a minimum highest intensity of 150,000, and a *m/z* tolerance of 0.001 *m/z* or 5 ppm. Deconvolution was performed using the Wavelets (ADAP) module, with *m/z* center calculation using median, and ranges for MS2 scan pairing of 0.01 Da and 0.4 min. The isotopes were grouped with a *m/z* tolerance of 0.001 *m/z* or 5 ppm and RT tolerance of 0.1 min. Peaks were aligned with a *m/z* tolerance of 0.001 *m/z* or 5 ppm and RT tolerance of 0.1 min, with 75% weight given to *m/z* and 25% to RT. Finally, peaks were filtered with a minimum of fifteen peaks in a row, and the same RT and peak duration ranges as previously applied. The feature quantification table (.csv) and aggregated MS2 masses list (.mgf) were exported (no merging of MS/MS and filter rows: ALL) for further analysis.

### Feature-based molecular networking using GNPS and compound annotation

A molecular network was created with the feature-based molecular networking workflow (https://ccms-ucsd.github.io/GNPSDocumentation/featurebasedmolecularnetworking/) [92] on the GNPS website (http://gnps.ucsd.edu) [93] by uploading the aggregated MS2 mass list. The data was filtered by removing all MS/MS fragment ions within +/- 17 Da of the precursor *m/z*. MS/MS spectra were window filtered by choosing only the top 6 fragment ions in the +/- 50Da window throughout the spectrum. The precursor ion mass tolerance was set to 0.02 Da and a MS/MS fragment ion tolerance of 0.02 Da. A network was then created where edges were filtered to have a cosine score above 0.7 and more than 4 matched peaks. Further, edges between two nodes were kept in the network if and only if each of the nodes appeared in each other’s respective top 10 most similar nodes. Finally, the maximum size of a molecular family was set to 100, and the lowest scoring edges were removed from molecular families until the molecular family size was below this threshold. The spectra in the network were then searched against GNPS’ spectral libraries. The library spectra were filtered in the same manner as the input data. All matches kept between network spectra and library spectra were required to have a score above 0.7 and at least 4 matched peaks. The .graphml network file was then visualized using Cytoscape v3.7.2 [94] where individual sample data and metadata were locally plotted (per sample and metadata sample group relative intensities). To enhance annotation of potential compounds of interest using the mzCloud spectral library (Thermo Fisher Scientific), .raw files were also preprocessed using Compound Discoverer 2.1 (CD2.1) SP1 software (Thermo Fisher Scientific). Details regarding the settings are provided in Document S2. GNPS and Compound Discoverer (annotation reported when above mzCloud 80% confidence in identity or similarity search) offer annotations with a level 2 confidence according to the Metabolomics Standards Initiative (i.e. putative annotation) [62,63]. To summarize and further enhance chemical structural information within the molecular network, substructure information (https://ccms-ucsd.github.io/GNPSDocumentation/ms2lda/) [95], information from *in silico* structure annotations from Network Annotation Propagation [96] and Dereplicator [97] were incorporated using the GNPS MolNetEnhancer workflow (https://ccms-ucsd.github.io/GNPSDocumentation/molnetenhancer/) [61] with chemical class annotations retrieved from the ClassyFire chemical ontology [98]. When no chemical structural information could be retrieved through the above searches, the MS/MS spectra were additionally searched via MASST [99] and SIRIUS+CSI:FingerID [100–102]. MASST allows to query a single MS/MS spectrum across all public GNPS datasets giving an idea of the type of samples or matrices where the same MS/MS spectrum has been detected [99]. SIRIUS+CSI:FingerID uses deep learning algorithms to predict the molecular and structural formula of a molecule from MS/MS spectra [100–103].

### Contamination filtering and further data curation

Using a Kendrick Mass Filter, we explored the compositionality of our data to assess the potential presence of undesired chemical background [104]. Out of the 4,360 features obtained through MZmine preprocessing, more than 1,100 possessed repeat units typical of polyethylene glycol (PEG) and polypropylene glycol (PPG). Filtering of PEG followed by filtering of PPG was performed using a Kendrick Mass Filter [104] with the following parameters: number of observed signals = 5, Kendrick mass defect = 0.01, and fraction base = 1. All Jupyter notebooks used are publicly available on GitHub (https://github.com/SSI-Metabolomics/Autism_SupplementaryMaterial).

Of the 3,253 remaining features, we further excluded those with a maximum intensity in paper blanks / maximum intensity in samples ratio ≥ 0.2, as well as features with 80% or more gaps (i.e. missing value) in cases and/or in controls (1,975 features filtered).

### Data visualization and outlier handling

To detect overall patterns in our data, we performed PCA, which allowed us to assess the consistency of repeated pool injections (i.e. repeated injections of the same pooled samples should cluster in PCA). When performing these calculations on our “raw” unfiltered feature table (4,360 features), seven samples were revealed as clear outliers, of which two controls and five cases. After contamination filtering and data curation (1,281 features), six outliers remained since one outlier (control) was due to PEG contamination. Among the investigated potential explanations for these outliers, no pattern was found when looking at: position on the plate layout, potential RT shift impairing the alignment, and metadata. However, targeted analysis of labeled internal standards and unlabeled endogenous homologs showed that significant (but unexplained) errors occurred during LC-MS/MS acquisition, with many undetectable compounds (TraceFinder 4.1 Clinical Research, Thermo Fischer Scientific) (Figure S1). A heatmap representation of the data (1,281 features) using MetaboAnalyst 4.0 [105] confirmed the six outliers with very low intensities (Figure S1). Therefore, we decided to exclude these outliers from further statistical analyses.

### Statistics

Using the filtered feature table, we calculated pair-wise Euclidean distances (1,281 features, 68 samples) and performed Permutational Multivariate Analyses of Variance (PERMANOVAs) [106] to assess how much of the variance in the data is explained by a certain variable in the metadata. We investigated the following variables: ASD (yes/no), ASD subtype, gender, birthweight, gestational age, age at sampling, month of birth, and injection order. The Adonis R^2^ value indicates to what extent the variance is explained by the tested variable. Significance threshold was set at 0.05. Calculations were performed using the ggplot2, ggfortify, ggsci, rlang, viridis and vegan packages in R software v3.6.1 [107].

Finally, the curated dataset (1,281 features, 68 samples, unpaired samples) was processed using MetaboAnalyst 4.0 [105] and R v3.6.1. We ran a fold-change analysis with a threshold of 2 (case/control or control/case). We then excluded features with more than 50% missing values and replaced the remaining missing values by a small value (half the minimum positive value in the original data). We further filtered non-informative near-constant features based on interquantile range and applied a glog transformation and Pareto scaling. We performed t-tests and Wilcoxon rank-sum tests with FDR correction for multiple comparisons. We could not reliably use the Partial Least Squares Discriminant Analysis (negative Q2 in cross validation). All Jupyter notebooks used for statistical analysis are publicly available on GitHub (https://github.com/SSI-Metabolomics/Autism_SupplementaryMaterial).

## 5. Conclusions

This is the first untargeted metabolomics study assessing the potential of biobanked newborn DBS samples in ASD research. The development of biobanks and reuse of systematically collected DBS samples for research purposes in connection with registry data represent many new opportunities to study the physiopathology and early signs of diseases, with extraordinary impacts in prevention, diagnosis and treatment strategies. We showed that untargeted metabolomics on DBS samples offers a wide and relevant coverage of metabolites for the study of ASD and that the new processing tools used in our method largely expand the interpretability of such complex data.

## Supporting information

DocumentS1_MZmine_batchProcessing

DocumentS2_CompoundDiscoverer2.1_workflow

FigureS1_Analysis_Outliers

TableS1_Subjects_characteristics_details

TableS2_Annotated_compounds

TableS3_PERMANOVAs_values

TableS4_Compounds_inspected

TableS5_Fragmentation_profiles

TableS6_Compounds_reported_literature

TableS7_StandardizedMethodDescription

## Data Availability

The datasets generated and/or analyzed during the current study are not publicly available due to the risk of compromising individual privacy but are available from the corresponding author on reasonable request and provided that an appropriate collaboration agreement can be agreed upon.

## Supplementary Materials

The following are available online at www.mdpi.com/xxx/s1, Table S1: Subjects characteristics in details, Table S2: All features including annotated compounds. Out of the 4360 features detected, 150 could be annotated by GNPS library matching (annotation level 2) or in-house TraceFinder library (annotation level 1) and an additional 859 by MolNetEnhancer (annotation level 3). Table S3: PERMANOVAs Adonis R2 values and p-values calculated without outliers (68 samples), Table S4: Full list of compounds with t-test p value <0.01 without FDR correction and/or with Wilcoxon rank-sum test p value <0.01 without FDR correction and/or case/control fold change value <0.5 or >2, Table S5: Fragmentation profiles of the two unknown features to be monitored in future studies as well as methacholine as shown in Table 2, Table S6: Full list of compounds reported in the literature as involved in ASD and considered in this study. Table S7: Standardized reporting of untargeted metabolomics LC-MS/MS method according to [87] containing the list of internal standards from the Neobase Non-derivatized MSMS kit and their concentration in the extraction buffer, Document S1: MZmine batch .xml file used to preprocess the raw data, Document S2: Compound Discoverer 2.1 preprocessing workflow settings, Figure S1: Targeted analysis of outliers using TraceFinder (IS and unlabeled homologs) and heatmap of untargeted analysis,

## Author Contributions

Conceptualization, J.C. and A.S.C..; methodology, J.C. and S.S.L.; software, J.C. and M.E.; validation, J.C. and M.E.; formal analysis, J.C.; investigation, J.C.; resources, A.S.C.; data curation, J.C.; writing— original draft preparation, J.C.; writing—review and editing, J.C., M.E., S.S.L. and A.S.C.; visualization, J.C.; supervision, A.S.C.; project administration, J.C.; funding acquisition, D.M.H. All authors have read and agreed to the published version of the manuscript.

## Acknowledgments

We thank Anders Björkbom for his major contribution to the LC-MS/MS method development and Marie Bækvad-Hansen for her help in curating data.

## References

1. CDC Data and Statistics on Autism Spectrum Disorder | CDC Available online: https://www.cdc.gov/ncbddd/autism/data.html (accessed on Jun 18, 2019).

2. Randall, M., Egberts, K.J., Samtani, A., Scholten, R.J., Hooft, L., Livingstone, N., Sterling-Levis, K., Woolfenden, S., Williams, K. Diagnostic tests for autism spectrum disorder (ASD) in preschool children. Cochrane Database Syst. Rev. 2018, doi:10.1002/14651858.CD009044.pub2.

3. World Health Organization The ICD-10 classification of mental and behavioural disorders: diagnostic criteria for research; World Health Organization: Geneva, 1993; ISBN 978-92-4-154455-9.

4. Adam, D. Mental health: On the spectrum. Nat. News 2013, 496, 416, doi:10.1038/496416a.

5. Frances, A. ICD, DSM and The Tower of Babel. Aust. N. Z. J. Psychiatry 2014, 48, 371–373, doi:10.1177/0004867414526792.

6. Bejarano-Martín, Á., Canal-Bedia, R., Magán-Maganto, M., Fernández-Álvarez, C., Cilleros-Martín, M.V., Sánchez-Gómez, M.C., García-Primo, P., Rose-Sweeney, M., Boilson, A., Linertová, R., et al. Early Detection, Diagnosis and Intervention Services for Young Children with Autism Spectrum Disorder in the European Union (ASDEU): Family and Professional Perspectives. J. Autism Dev. Disord. 2019, doi:10.1007/s10803-019-04253-0.

7. Shen, L., Liu, X., Zhang, H., Lin, J., Feng, C., Iqbal, J. Biomarkers in autism spectrum disorders: Current progress. Clin. Chim. Acta Int. J. Clin. Chem. 2019, 502, 41–54, doi:10.1016/j.cca.2019.12.009.

8. Bai, D., Yip, B.H.K., Windham, G.C., Sourander, A., Francis, R., Yoffe, R., Glasson, E., Mahjani, B., Suominen, A., Leonard, H., et al. Association of Genetic and Environmental Factors With Autism in a 5-Country Cohort. JAMA Psychiatry 2019, 76, 1035–1043, doi:10.1001/jamapsychiatry.2019.1411.

9. Grove, J., Ripke, S., Als, T.D., Mattheisen, M., Walters, R.K., Won, H., Pallesen, J., Agerbo, E., Andreassen, O.A., Anney, R., et al. Identification of common genetic risk variants for autism spectrum disorder. Nat. Genet. 2019, 51, 431–444, doi:10.1038/s41588-019-0344-8.

10. Newschaffer, C.J., Croen, L.A., Daniels, J., Giarelli, E., Grether, J.K., Levy, S.E., Mandell, D.S., Miller, L.A., Pinto-Martin, J., Reaven, J., et al. The epidemiology of autism spectrum disorders. Annu. Rev. Public Health 2007, 28, 235–258, doi:10.1146/annurev.publhealth.28.021406.144007.

11. Hannon, E., Schendel, D., Ladd-Acosta, C., Grove, J., Hansen, C.S., Andrews, S.V., Hougaard, D.M., Bresnahan, M., Mors, O., Hollegaard, M.V., et al. Elevated polygenic burden for autism is associated with differential DNA methylation at birth. Genome Med. 2018, 10, doi:10.1186/s13073-018-0527-4.

12. Braam, W., Ehrhart, F., Maas, A.P.H.M., Smits, M.G., Curfs, L. Low maternal melatonin level increases autism spectrum disorder risk in children. Res. Dev. Disabil. 2018, 82, 79–89, doi:10.1016/j.ridd.2018.02.017.

13. Abbott, P.W., Gumusoglu, S.B., Bittle, J., Beversdorf, D.Q., Stevens, H.E. Prenatal stress and genetic risk: How prenatal stress interacts with genetics to alter risk for psychiatric illness. Psychoneuroendocrinology 2018, 90, 9–21, doi:10.1016/j.psyneuen.2018.01.019.

14. Fine, R., Zhang, J., Stevens, H.E. Prenatal stress and inhibitory neuron systems: implications for neuropsychiatric disorders. Mol. Psychiatry 2014, 19, 641–651, doi:10.1038/mp.2014.35.

15. Croen, L.A., Qian, Y., Ashwood, P., Zerbo, O., Schendel, D., Pinto-Martin, J., Fallin, M.D., Levy, S., Schieve, L.A., Yeargin-Allsopp, M., et al. Infection and Fever in Pregnancy and Autism Spectrum Disorders: Findings from the Study to Explore Early Development. Autism Res. 2019, 12, 1551–1561, doi:10.1002/aur.2175.

16. Kuzniewicz, M.W., Wi, S., Qian, Y., Walsh, E.M., Armstrong, M.A., Croen, L.A. Prevalence and Neonatal Factors Associated with Autism Spectrum Disorders in Preterm Infants. J. Pediatr. 2014, 164, 20–25, doi:10.1016/j.jpeds.2013.09.021.

17. Apgar, V. A proposal for a new method of evaluation of the newborn infant. Curr. Res. Anesth. Analg. 1953, 32, 260–267.

18. Modabbernia, A., Sandin, S., Gross, R., Leonard, H., Gissler, M., Parner, E.T., Francis, R., Carter, K., Bresnahan, M., Schendel, D., et al. Apgar score and risk of autism. Eur. J. Epidemiol. 2019, 34, 105–114, doi:10.1007/s10654-018-0445-1.

19. Alberti, A., Pirrone, P., Elia, M., Waring, R.H., Romano, C. Sulphation deficit in “low-functioning” autistic children: a pilot study. Biol. Psychiatry 1999, 46, 420–424, doi:10.1016/S0006-3223(98)00337-0.

20. Chaidez, V., Hansen, R.L., Hertz-Picciotto, I. Gastrointestinal problems in children with autism, developmental delays or typical development. J. Autism Dev. Disord. 2014, 44, 1117–1127, doi:10.1007/s10803-013-1973-x.

21. Krajmalnik-Brown, R., Lozupone, C., Kang, D.-W., Adams, J.B. Gut bacteria in children with autism spectrum disorders: challenges and promise of studying how a complex community influences a complex disease. Microb. Ecol. Health Dis. 2015, 26, doi:10.3402/mehd.v26.26914.

22. White, J.F. Intestinal pathophysiology in autism. Exp. Biol. Med. Maywood NJ 2003, 228, 639–649, doi:10.1177/153537020322800601.

23. Cerdó, T., Diéguez, E., Campoy, C. Early nutrition and gut microbiome: interrelationship between bacterial metabolism, immune system, brain structure, and neurodevelopment. Am. J. Physiol.-Endocrinol. Metab. 2019, 317, E617–E630, doi:10.1152/ajpendo.00188.2019.

24. Wang, S., Harvey, L., Martin, R., van der Beek, E.M., Knol, J., Cryan, J.F., Renes, I.B. Targeting the gut microbiota to influence brain development and function in early life. Neurosci. Biobehav. Rev. 2018, 95, 191–201, doi:10.1016/j.neubiorev.2018.09.002.

25. De Angelis, M., Francavilla, R., Piccolo, M., De Giacomo, A., Gobbetti, M. Autism spectrum disorders and intestinal microbiota. Gut Microbes 2015, 6, 207–213, doi:10.1080/19490976.2015.1035855.

26. Sharon, G., Cruz, N.J., Kang, D.-W., Gandal, M.J., Wang, B., Kim, Y.-M., Zink, E.M., Casey, C.P., Taylor, B.C., Lane, C.J., et al. Human Gut Microbiota from Autism Spectrum Disorder Promote Behavioral Symptoms in Mice. Cell 2019, 177, 1600-1618.e17, doi:10.1016/j.cell.2019.05.004.

27. Kelly, R.S., Boulin, A., Laranjo, N., Lee-Sarwar, K., Chu, S.H., Yadama, A.P., Carey, V., Litonjua, A.A., Lasky-Su, J., Weiss, S.T. Metabolomics and Communication Skills Development in Children; Evidence from the Ages and Stages Questionnaire. Metabolites 2019, 9, 42, doi:10.3390/metabo9030042.

28. Sgritta, M., Dooling, S.W., Buffington, S.A., Momin, E.N., Francis, M.B., Britton, R.A., Costa-Mattioli, M. Mechanisms Underlying Microbial-Mediated Changes in Social Behavior in Mouse Models of Autism Spectrum Disorder. Neuron 2019, 101, 246-259.e6, doi:10.1016/j.neuron.2018.11.018.

29. Adams, J.B., Borody, T.J., Kang, D.-W., Khoruts, A., Krajmalnik-Brown, R., Sadowsky, M.J. Microbiota transplant therapy and autism: lessons for the clinic. Expert Rev. Gastroenterol. Hepatol. 2019, 13, 1033–1037, doi:10.1080/17474124.2019.1687293.

30. Adams, J.B., Vargason, T., Kang, D.-W., Krajmalnik-Brown, R., Hahn, J. Multivariate Analysis of Plasma Metabolites in Children with Autism Spectrum Disorder and Gastrointestinal Symptoms Before and After Microbiota Transfer Therapy. Processes 2019, 7, 806, doi:10.3390/pr7110806.

31. Borre, Y.E., O’Keeffe, G.W., Clarke, G., Stanton, C., Dinan, T.G., Cryan, J.F. Microbiota and neurodevelopmental windows: implications for brain disorders. Trends Mol. Med. 2014, 20, 509–518, doi:10.1016/j.molmed.2014.05.002.

32. Magistris, L. de; Familiari, V., Pascotto, A., Sapone, A., Frolli, A., Iardino, P., Carteni, M., Rosa, M.D., Francavilla, R., Riegler, G., et al. Alterations of the Intestinal Barrier in Patients With Autism Spectrum Disorders and in Their First-degree Relatives. J. Pediatr. Gastroenterol. Nutr. 2010, 51, 418–424, doi:10.1097/MPG.0b013e3181dcc4a5.

33. Aldred, S., Moore, K.M., Fitzgerald, M., Waring, R.H. Plasma Amino Acid Levels in Children with Autism and Their Families. J. Autism Dev. Disord. 2003, 33, 93–97, doi:10.1023/A:1022238706604.

34. Evans, C., Dunstan, R.H., Rothkirch, T., Roberts, T.K., Reichelt, K.L., Cosford, R., Deed, G., Ellis, L.B., Sparkes, D.L. Altered amino acid excretion in children with autism. Nutr. Neurosci. 2008, 11, 9–17, doi:10.1179/147683008X301360.

35. De Angelis, M., Piccolo, M., Vannini, L., Siragusa, S., De Giacomo, A., Serrazzanetti, D.I., Cristofori, F., Guerzoni, M.E., Gobbetti, M., Francavilla, R. Fecal Microbiota and Metabolome of Children with Autism and Pervasive Developmental Disorder Not Otherwise Specified. Plos One 2013, 8, e76993, doi:10.1371/journal.pone.0076993.

36. Kang, D.-W., Ilhan, Z.E., Isern, N.G., Hoyt, D.W., Howsmon, D.P., Shaffer, M., Lozupone, C.A., Hahn, J., Adams, J.B., Krajmalnik-Brown, R. Differences in fecal microbial metabolites and microbiota of children with autism spectrum disorders. Anaerobe 2018, 49, 121–131, doi:10.1016/j.anaerobe.2017.12.007.

37. Naushad, S.M., Jain, J.M.N., Prasad, C.K., Naik, U., Akella, R.R.D. Autistic children exhibit distinct plasma amino acid profile. Indian J. Biochem. Biophys. 2013, 50, 474–478.

38. Barone, R., Alaimo, S., Messina, M., Pulvirenti, A., Bastin, J., Group, M.-A., Ferro, A., Frye, R.E., Rizzo, R., Fiumara, A., et al. A Subset of Patients With Autism Spectrum Disorders Show a Distinctive Metabolic Profile by Dried Blood Spot Analyses. Front. Psychiatry 2018, 9, doi:10.3389/fpsyt.2018.00636.

39. Anwar, A., Abruzzo, P.M., Pasha, S., Rajpoot, K., Bolotta, A., Ghezzo, A., Marini, M., Posar, A., Visconti, P., Thornalley, P.J., et al. Advanced glycation endproducts, dityrosine and arginine transporter dysfunction in autism - a source of biomarkers for clinical diagnosis. Mol. Autism 2018, 9, 3, doi:10.1186/s13229-017-0183-3.

40. Smith, A.M., King, J.J., West, P.R., Ludwig, M.A., Donley, E.L.R., Burrier, R.E., Amaral, D.G. Amino Acid Dysregulation Metabotypes: Potential Biomarkers for Diagnosis and Individualized Treatment for Subtypes of Autism Spectrum Disorder. Biol. Psychiatry 2019, 85, 345–354, doi:10.1016/j.biopsych.2018.08.016.

41. West, P.R., Amaral, D.G., Bais, P., Smith, A.M., Egnash, L.A., Ross, M.E., Palmer, J.A., Fontaine, B.R., Conard, K.R., Corbett, B.A., et al. Metabolomics as a Tool for Discovery of Biomarkers of Autism Spectrum Disorder in the Blood Plasma of Children. PLoS ONE 2014, 9, doi:10.1371/journal.pone.0112445.

42. Rangel-Huerta, O.D., Gomez-Fernández, A., de la Torre-Aguilar, M.J., Gil, A., Perez-Navero, J.L., Flores-Rojas, K., Martín-Borreguero, P., Gil-Campos, M. Metabolic profiling in children with autism spectrum disorder with and without mental regression: preliminary results from a cross-sectional case-control study. Metabolomics Off. J. Metabolomic Soc. 2019, 15, 99, doi:10.1007/s11306-019-1562-x.

43. Bitar, T., Mavel, S., Emond, P., Nadal-Desbarats, L., Lefèvre, A., Mattar, H., Soufia, M., Blasco, H., Vourc’h, P., Hleihel, W., et al. Identification of metabolic pathway disturbances using multimodal metabolomics in autistic disorders in a Middle Eastern population. J. Pharm. Biomed. Anal. 2018, 152, 57–65, doi:10.1016/j.jpba.2018.01.007.

44. Delaye, J.-B., Patin, F., Lagrue, E., Le Tilly, O., Bruno, C., Vuillaume, M.-L., Raynaud, M., Benz-De Bretagne, I., Laumonnier, F., Vourc’h, P., et al. Post hoc analysis of plasma amino acid profiles: towards a specific pattern in autism spectrum disorder and intellectual disability. Ann. Clin. Biochem. 2018, 55, 543–552, doi:10.1177/0004563218760351.

45. Kuwabara, H., Yamasue, H., Koike, S., Inoue, H., Kawakubo, Y., Kuroda, M., Takano, Y., Iwashiro, N., Natsubori, T., Aoki, Y., et al. Altered Metabolites in the Plasma of Autism Spectrum Disorder: A Capillary Electrophoresis Time-of-Flight Mass Spectroscopy Study. PLoS ONE 2013, 8, doi:10.1371/journal.pone.0073814.

46. Smith, A.M., Natowicz, M.R., Braas, D., Ludwig, M.A., Ney, D.M., Donley, E.L.R., Burrier, R.E., Amaral, D.G. A Metabolomics Approach to Screening for Autism Risk in the Children’s Autism Metabolome Project. Autism Res. 2020, 13, 1270–1285, doi:10.1002/aur.2330.

47. Retey, J. The Urocanase Story: A Novel Role of NAD+ as Electrophile. Arch. Biochem. Biophys. 1994, 314, 1–16, doi:10.1006/abbi.1994.1405.

48. Shaw, W. Increased urinary excretion of a 3-(3-hydroxyphenyl)-3-hydroxypropionic acid (HPHPA), an abnormal phenylalanine metabolite of Clostridia spp. in the gastrointestinal tract, in urine samples from patients with autism and schizophrenia. Nutr. Neurosci. 2010, 13, 135–143, doi:10.1179/147683010X12611460763968.

49. Mavel, S., Nadal-Desbarats, L., Blasco, H., Bonnet-Brilhault, F., Barthélémy, C., Montigny, F., Sarda, P., Laumonnier, F., Vourc’h, P., Andres, C.R., et al. 1H-13C NMR-based urine metabolic profiling in autism spectrum disorders. Talanta 2013, 114, 95–102, doi:10.1016/j.talanta.2013.03.064.

50. Yap, I.K.S., Angley, M., Veselkov, K.A., Holmes, E., Lindon, J.C., Nicholson, J.K. Urinary metabolic phenotyping differentiates children with autism from their unaffected siblings and age-matched controls. J. Proteome Res. 2010, 9, 2996–3004, doi:10.1021/pr901188e.

51. Ming, X., Stein, T.P., Barnes, V., Rhodes, N., Guo, L. Metabolic perturbance in autism spectrum disorders: a metabolomics study. J. Proteome Res. 2012, 11, 5856–5862, doi:10.1021/pr300910n.

52. Liu, A., Zhou, W., Qu, L., He, F., Wang, H., Wang, Y., Cai, C., Li, X., Zhou, W., Wang, M. Altered Urinary Amino Acids in Children With Autism Spectrum Disorders. Front. Cell. Neurosci. 2019, 13, doi:10.3389/fncel.2019.00007.

53. Kałużna-Czaplińska, J. Noninvasive urinary organic acids test to assess biochemical and nutritional individuality in autistic children. Clin. Biochem. 2011, 44, 686–691, doi:10.1016/j.clinbiochem.2011.01.015.

54. Lussu, M., Noto, A., Masili, A., Rinaldi, A.C., Dessì, A., Angelis, M.D., Giacomo, A.D., Fanos, V., Atzori, L., Francavilla, R. The urinary 1H-NMR metabolomics profile of an italian autistic children population and their unaffected siblings. Autism Res. 2017, 10, 1058–1066, doi:10.1002/aur.1748.

55. Chen, Q., Qiao, Y., Xu, X., You, X., Tao, Y. Urine Organic Acids as Potential Biomarkers for Autism-Spectrum Disorder in Chinese Children. Front. Cell. Neurosci. 2019, 13, doi:10.3389/fncel.2019.00150.

56. Emond, P., Mavel, S., Aïdoud, N., Nadal-Desbarats, L., Montigny, F., Bonnet-Brilhault, F., Barthélémy, C., Merten, M., Sarda, P., Laumonnier, F., et al. GC-MS-based urine metabolic profiling of autism spectrum disorders. Anal. Bioanal. Chem. 2013, 405, 5291–5300, doi:10.1007/s00216-013-6934-x.

57. Noto, A., Fanos, V., Barberini, L., Grapov, D., Fattuoni, C., Zaffanello, M., Casanova, A., Fenu, G., Giacomo, A.D., Angelis, M.D., et al. The urinary metabolomics profile of an Italian autistic children population and their unaffected siblings. J. Matern. Fetal Neonatal Med. 2014, 27, 46–52, doi:10.3109/14767058.2014.954784.

58. Gevi, F., Zolla, L., Gabriele, S., Persico, A.M. Urinary metabolomics of young Italian autistic children supports abnormal tryptophan and purine metabolism. Mol. Autism 2016, 7, 47, doi:10.1186/s13229-016-0109-5.

59. Wang, M., Wan, J., Rong, H., He, F., Wang, H., Zhou, J., Cai, C., Wang, Y., Xu, R., Yin, Z., et al. Alterations in Gut Glutamate Metabolism Associated with Changes in Gut Microbiota Composition in Children with Autism Spectrum Disorder. mSystems 2019, 4, doi:10.1128/mSystems.00321-18.

60. Nørgaard-Pedersen, B., Hougaard, D.M. Storage policies and use of the Danish Newborn Screening Biobank. J. Inherit. Metab. Dis. 2007, 30, 530–536, doi:10.1007/s10545-007-0631-x.

61. Ernst, M., Kang, K.B., Caraballo-Rodríguez, A.M., Nothias, L.-F., Wandy, J., Chen, C., Wang, M., Rogers, S., Medema, M.H., Dorrestein, P.C., et al. MolNetEnhancer: Enhanced Molecular Networks by Integrating Metabolome Mining and Annotation Tools. Metabolites 2019, 9, 144, doi:10.3390/metabo9070144.

62. Schrimpe-Rutledge, A.C., Codreanu, S.G., Sherrod, S.D., McLean, J.A. Untargeted Metabolomics Strategies—Challenges and Emerging Directions. J. Am. Soc. Mass Spectrom. 2016, 27, 1897–1905, doi:10.1007/s13361-016-1469-y.

63. Sumner, L.W., Amberg, A., Barrett, D., Beale, M.H., Beger, R., Daykin, C.A., Fan, T.W.-M., Fiehn, O., Goodacre, R., Griffin, J.L., et al. Proposed minimum reporting standards for chemical analysis: Chemical Analysis Working Group (CAWG) Metabolomics Standards Initiative (MSI). Metabolomics 2007, 3, 211–221, doi:10.1007/s11306-007-0082-2.

64. Glinton, K.E., Elsea, S.H. Untargeted Metabolomics for Autism Spectrum Disorders: Current Status and Future Directions. Front. Psychiatry 2019, 10, doi:10.3389/fpsyt.2019.00647.

65. Mizejewski, G.J., Lindau-Shepard, B., Pass, K.A. Newborn screening for autism: in search of candidate biomarkers. Biomark. Med. 2013, 7, 247–260, doi:10.2217/bmm.12.108.

66. Lee, M., Martin-Ruiz, C., Graham, A., Court, J., Jaros, E., Perry, R., Iversen, P., Bauman, M., Perry, E. Nicotinic receptor abnormalities in the cerebellar cortex in autism. Brain J. Neurol. 2002, 125, 1483–1495, doi:10.1093/brain/awf160.

67. Janecka, M., Kodesh, A., Levine, S.Z., Lusskin, S.I., Viktorin, A., Rahman, R., Buxbaum, J.D., Schlessinger, A., Sandin, S., Reichenberg, A. Association of Autism Spectrum Disorder With Prenatal Exposure to Medication Affecting Neurotransmitter Systems. JAMA Psychiatry 2018, 75, 1217–1224, doi:10.1001/jamapsychiatry.2018.2728.

68. Saigal, S., Doyle, L.W. An overview of mortality and sequelae of preterm birth from infancy to adulthood. The Lancet 2008, 371, 261–269, doi:10.1016/S0140-6736(08)60136-1.

69. Gil, A.M., Duarte, D. Biofluid Metabolomics in Preterm Birth Research. Reprod. Sci. 2018, 25, 967–977, doi:10.1177/1933719118756748.

70. Ernst, M., Rogers, S., Lausten-Thomsen, U., Björkbom, A., Laursen, S.S., Courraud, J., Børglum, A., Nordentoft, M., Werge, T., Mortensen, P.B., et al. Gestational age-dependent development of the neonatal metabolome. Pediatr. Res. 2020, 1–9, doi:10.1038/s41390-020-01149-z.

71. Dionisi-Vici, C., Deodato, F., Roschinger, W., Rhead, W., Wilcken, B. “Classical” organic acidurias, propionic aciduria, methylmalonic aciduria and isovaleric aciduria: Long-term outcome and effects of expanded newborn screening using tandem mass spectrometry. J. Inherit. Metab. Dis. 2006, 29, 383–389, doi:10.1007/s10545-006-0278-z.

72. Pedersen, C.B., Bybjerg-Grauholm, J., Pedersen, M.G., Grove, J., Agerbo, E., Bækvad-Hansen, M., Poulsen, J.B., Hansen, C.S., McGrath, J.J., Als, T.D., et al. The iPSYCH2012 case–cohort sample: new directions for unravelling genetic and environmental architectures of severe mental disorders. Mol. Psychiatry 2018, 23, 6–14, doi:10.1038/mp.2017.196.

73. Milani, C., Duranti, S., Bottacini, F., Casey, E., Turroni, F., Mahony, J., Belzer, C., Delgado Palacio, S., Arboleya Montes, S., Mancabelli, L., et al. The First Microbial Colonizers of the Human Gut: Composition, Activities, and Health Implications of the Infant Gut Microbiota. Microbiol. Mol. Biol. Rev. MMBR 2017, 81, doi:10.1128/MMBR.00036-17.

74. Backhed, F., Roswall, J., Peng, Y., Feng, Q., Jia, H., Kovatcheva-Datchary, P., Li, Y., Xia, Y., Xie, H., Zhong, H., et al. Dynamics and Stabilization of the Human Gut Microbiome during the First Year of Life. Cell Host Microbe 2015, 17, 690–703, doi:10.1016/j.chom.2015.04.004.

75. Rosa, P.S.L., Warner, B.B., Zhou, Y., Weinstock, G.M., Sodergren, E., Hall-Moore, C.M., Stevens, H.J., Bennett, W.E., Shaikh, N., Linneman, L.A., et al. Patterned progression of bacterial populations in the premature infant gut. Proc. Natl. Acad. Sci. 2014, 111, 12522–12527, doi:10.1073/pnas.1409497111.

76. Wilmanski, T., Rappaport, N., Earls, J.C., Magis, A.T., Manor, O., Lovejoy, J., Omenn, G.S., Hood, L., Gibbons, S.M., Price, N.D. Blood metabolome predicts gut microbiome α-diversity in humans. Nat. Biotechnol. 2019, 37, 1217–1228, doi:10.1038/s41587-019-0233-9.

77. Meng, D., Sommella, E., Salviati, E., Campiglia, P., Ganguli, K., Djebali, K., Zhu, W., Walker, W.A. Indole-3-lactic acid, a metabolite of tryptophan, secreted by Bifidobacterium longum subspecies infantis is anti-inflammatory in the immature intestine. Pediatr. Res. 2020, 1–9, doi:10.1038/s41390-019-0740-x.

78. Laursen, M.F., Sakanaka, M., Burg, N. von; Andersen, D., Mörbe, U., Rivollier, A., Pekmez, C.T., Moll, J.M., Michaelsen, K.F., Mølgaard, C., et al. Breastmilk-promoted bifidobacteria produce aromatic lactic acids in the infant gut. bioRxiv 2020, 2020.01.22.914994, doi:10.1101/2020.01.22.914994.

79. Keller, M.C., Fredrickson, B.L., Ybarra, O., Cote, S., Johnson, K., Mikels, J., Conway, A., Wager, T. A warm heart and a clear head -The contingent effects of weather on mood and cognition. Psychol. Sci. 2005, 16, 724–731, doi:10.1111/j.1467-9280.2005.01602.x.

80. D’Amato, G., Holgate, S.T., Pawankar, R., Ledford, D.K., Cecchi, L., Al-Ahmad, M., Al-Enezi, F., Al-Muhsen, S., Ansotegui, I., Baena-Cagnani, C.E., et al. Meteorological conditions, climate change, new emerging factors, and asthma and related allergic disorders. A statement of the World Allergy Organization. World Allergy Organ. J. 2015, 8, UNSP 25, doi:10.1186/s40413-015-0073-0.

81. Ruoppolo, M., Scolamiero, E., Caterino, M., Mirisola, V., Franconi, F., Campesi, I. Female and male human babies have distinct blood metabolomic patterns. Mol. Biosyst. 2015, 11, 2483–2492, doi:10.1039/c5mb00297d.

82. Diaz, S.O., Pinto, J., Barros, A.S., Morais, E., Duarte, D., Negrão, F., Pita, C., Almeida, M. do C., Carreira, I.M., Spraul, M., et al. Newborn Urinary Metabolic Signatures of Prematurity and Other Disorders: A Case Control Study. J. Proteome Res. 2016, 15, 311–325, doi:10.1021/acs.jproteome.5b00977.

83. Challis, J., Newnham, J., Petraglia, F., Yeganegi, M., Bocking, A. Fetal sex and preterm birth. Placenta 2013, 34, 95–99, doi:10.1016/j.placenta.2012.11.007.

84. Wilkin, T.J., Murphy, M.J. The gender insulin hypothesis: why girls are born lighter than boys, and the implications for insulin resistance. Int. J. Obes. 2006, 30, 1056–1061, doi:10.1038/sj.ijo.0803317.

85. Petrick, L., Edmands, W., Schiffman, C., Grigoryan, H., Perttula, K., Yano, Y., Dudoit, S., Whitehead, T., Metayer, C., Rappaport, S. An untargeted metabolomics method for archived newborn dried blood spots in epidemiologic studies. Metabolomics Off. J. Metabolomic Soc. 2017, 13, doi:10.1007/s11306-016-1153-z.

86. Folketinget Bekendtgørelse af lov om videnskabsetisk behandling af sundhedsvidenskabelige forskningsprojekter - (The Danish Parliament. Order of law on scientific treatment of health scientific research projects) Available online: https://www.retsinformation.dk/Forms/r0710.aspx?id=192671 (accessed on Oct 28, 2019).

87. Vogeser, M., Schuster, C., Rockwood, A.L. A proposal to standardize the description of LC–MS-based measurement methods in laboratory medicine. Clin. Mass Spectrom. 2019, 13, 36–38, doi:10.1016/j.clinms.2019.04.003.

88. Chambers, M.C., Maclean, B., Burke, R., Amodei, D., Ruderman, D.L., Neumann, S., Gatto, L., Fischer, B., Pratt, B., Egertson, J., et al. A cross-platform toolkit for mass spectrometry and proteomics. Nat. Biotechnol. 2012, 30, 918–920, doi:10.1038/nbt.2377.

89. Katajamaa, M., Miettinen, J., Oresic, M. MZmine: toolbox for processing and visualization of mass spectrometry based molecular profile data. Bioinforma. Oxf. Engl. 2006, 22, 634–636, doi:10.1093/bioinformatics/btk039.

90. Pluskal, T., Castillo, S., Villar-Briones, A., Orešič, M. MZmine 2: Modular framework for processing, visualizing, and analyzing mass spectrometry-based molecular profile data. BMC Bioinformatics 2010, 11, 395, doi:10.1186/1471-2105-11-395.

91. Myers, O.D., Sumner, S.J., Li, S., Barnes, S., Du, X. One Step Forward for Reducing False Positive and False Negative Compound Identifications from Mass Spectrometry Metabolomics Data: New Algorithms for Constructing Extracted Ion Chromatograms and Detecting Chromatographic Peaks. Anal. Chem. 2017, 89, 8696–8703, doi:10.1021/acs.analchem.7b00947.

92. Nothias, L.-F., Petras, D., Schmid, R., Dührkop, K., Rainer, J., Sarvepalli, A., Protsyuk, I., Ernst, M., Tsugawa, H., Fleischauer, M., et al. Feature-based molecular networking in the GNPS analysis environment. Nat. Methods 2020, 17, 905–908, doi:10.1038/s41592-020-0933-6.

93. Wang, M., Carver, J.J., Phelan, V.V., Sanchez, L.M., Garg, N., Peng, Y., Nguyen, D.D., Watrous, J., Kapono, C.A., Luzzatto-Knaan, T., et al. Sharing and community curation of mass spectrometry data with Global Natural Products Social Molecular Networking. Nat. Biotechnol. 2016, 34, 828–837, doi:10.1038/nbt.3597.

94. Shannon, P., Markiel, A., Ozier, O., Baliga, N.S., Wang, J.T., Ramage, D., Amin, N., Schwikowski, B., Ideker, T. Cytoscape: a software environment for integrated models of biomolecular interaction networks. Genome Res. 2003, 13, 2498–2504, doi:10.1101/gr.1239303.

95. Hooft, J.J.J. van der; Wandy, J., Barrett, M.P., Burgess, K.E.V., Rogers, S. Topic modeling for untargeted substructure exploration in metabolomics. Proc. Natl. Acad. Sci. 2016, 113, 13738–13743, doi:10.1073/pnas.1608041113.

96. Silva, R.R. da; Wang, M., Nothias, L.-F., Hooft, J.J.J. van der; Caraballo-Rodríguez, A.M., Fox, E., Balunas, M.J., Klassen, J.L., Lopes, N.P., Dorrestein, P.C. Propagating annotations of molecular networks using in silico fragmentation. PLOS Comput. Biol. 2018, 14, e1006089, doi:10.1371/journal.pcbi.1006089.

97. Mohimani, H., Gurevich, A., Mikheenko, A., Garg, N., Nothias, L.-F., Ninomiya, A., Takada, K., Dorrestein, P.C., Pevzner, P.A. Dereplication of peptidic natural products through database search of mass spectra. Nat. Chem. Biol. 2017, 13, 30–37, doi:10.1038/nchembio.2219.

98. Djoumbou Feunang, Y., Eisner, R., Knox, C., Chepelev, L., Hastings, J., Owen, G., Fahy, E., Steinbeck, C., Subramanian, S., Bolton, E., et al. ClassyFire: automated chemical classification with a comprehensive, computable taxonomy. J. Cheminformatics 2016, 8, 61, doi:10.1186/s13321-016-0174-y.

99. Wang, M., Jarmusch, A.K., Vargas, F., Aksenov, A.A., Gauglitz, J.M., Weldon, K., Petras, D., Silva, R. da; Quinn, R., Melnik, A.V., et al. Mass spectrometry searches using MASST. Nat. Biotechnol. 2020, 38, 23–26, doi:10.1038/s41587-019-0375-9.

100. Böcker, S., Dührkop, K. Fragmentation trees reloaded. J. Cheminformatics 2016, 8, 5, doi:10.1186/s13321-016-0116-8.

101. Dührkop, K., Shen, H., Meusel, M., Rousu, J., Böcker, S. Searching molecular structure databases with tandem mass spectra using CSI:FingerID. Proc. Natl. Acad. Sci. U. S. A. 2015, 112, 12580–12585, doi:10.1073/pnas.1509788112.

102. Dührkop, K., Fleischauer, M., Ludwig, M., Aksenov, A.A., Melnik, A.V., Meusel, M., Dorrestein, P.C., Rousu, J., Böcker, S. SIRIUS 4: a rapid tool for turning tandem mass spectra into metabolite structure information. Nat. Methods 2019, 16, 299–302, doi:10.1038/s41592-019-0344-8.

103. Shen, H., Dührkop, K., Böcker, S., Rousu, J. Metabolite identification through multiple kernel learning on fragmentation trees. Bioinforma. Oxf. Engl. 2014, 30, i157–164, doi:10.1093/bioinformatics/btu275.

104. da Silva, R.R., Vargas, F., Ernst, M., Nguyen, N.H., Bolleddu, S., del Rosario, K.K., Tsunoda, S.M., Dorrestein, P.C., Jarmusch, A.K. Computational Removal of Undesired Mass Spectral Features Possessing Repeat Units via a Kendrick Mass Filter. J. Am. Soc. Mass Spectrom. 2019, 30, 268–277, doi:10.1007/s13361-018-2069-9.

105. Chong, J., Soufan, O., Li, C., Caraus, I., Li, S., Bourque, G., Wishart, D.S., Xia, J. MetaboAnalyst 4.0: towards more transparent and integrative metabolomics analysis. Nucleic Acids Res. 2018, 46, W486–W494, doi:10.1093/nar/gky310.

106. Anderson, M.J. A new method for non-parametric multivariate analysis of variance. Austral Ecol. 2001, 26, 32–46, doi:10.1111/j.1442-9993.2001.01070.pp.x.

107. R Core Team R: a language and environment for statistical computing Available online: https://www.R-project.org/ (accessed on Nov 21, 2019).

