## Supplementary material for "Studying autism using untargeted metabolomics in newborn screening samples": DocumentS2_CompoundDiscoverer2.1_workflow

Search name:
20190718_Autism_UNPaired_v2.6_NoNorm_noGF_7sc_allAdd_mzClO_DBs_RT_0.1min_1E5_0.1min_DBS
Search description: Untargeted Metabolomics workflow: Retention time alignment, Component Detection, Grouping, Elemental Composition Prediction, Gap Filling, Hide chemical Background (using blanks), ID using mzCloud (needs MS/MS) and ChemSpider (using Formula); KEGG Pathway Mapping and Differential Analysis (ANOVA, adjusted p-values, fold change, CV, etc.)
Search date: 18-07-2019 18:15:51
Created with Discoverer version: 2.1.0.401

[Input Files (6)]
-->Select Spectra (7)

[Select Spectra (7)]
-->Align Retention Times (10)
-->Create Mass Trace (28)

[Align Retention Times (10)]
-->Detect Unknown Compounds (9)

[Detect Unknown Compounds (9)]
-->Group Unknown Compounds (8)

[Group Unknown Compounds (8)]
-->Search mzCloud (22)
-->Predict Compositions (16)
-->Search ChemSpider (23)
-->Search Mass Lists (27)
-->Mark Background Compounds (20)

[Predict Compositions (16)]
-->Search ChemSpider (23)

[Search mzCloud (22)]

[Search ChemSpider (23)]

[Search Mass Lists (27)]

[Mark Background Compounds (20)]

[Create Mass Trace (28)]

[Differential Analysis (17)]

[Descriptive Statistics (24)]

[Assign Compound Annotations (29)]

----------------------------------------------------------------
Processing node 6: Input Files
----------------------------------------------------------------
Input Data:
- File Name(s) (Hidden):

----------------------------------------------------------------
Processing node 7: Select Spectra
----------------------------------------------------------------
1. General Settings:
- Precursor Selection:  Use MS(n - 1) Precursor
- Use New Precursor Reevaluation:  True
- Use Isotope Pattern in Precursor Reevaluation:  True
- Store Chromatograms:  False

2. Spectrum Properties Filter:
- Lower RT Limit:  0
- Upper RT Limit:  15
- First Scan:  0
- Last Scan:  0
- Ignore Specified Scans:  (not specified)
- Lowest Charge State:  1
- Highest Charge State:  3
- Min. Precursor Mass:  70 Da
- Max. Precursor Mass:  1050 Da
- Total Intensity Threshold:  500000
- Minimum Peak Count:  1

3. Scan Event Filters:
- Mass Analyzer:  Is FTMS
- MS Order:  Any
- Activation Type:  Is HCD
- Min. Collision Energy:  0
- Max. Collision Energy:  1000
- Scan Type:  Any
- Polarity Mode:  Is +

4. Peak Filters:
- S/N Threshold (FT-only):  3

5. Replacements for Unrecognized Properties:
- Unrecognized Charge Replacements:  1
- Unrecognized Mass Analyzer Replacements:  FTMS
- Unrecognized MS Order Replacements:  MS2
- Unrecognized Activation Type Replacements:  HCD
- Unrecognized Polarity Replacements:  +
- Unrecognized MS Resolution@200 Replacements:  75000
- Unrecognized MSn Resolution@200 Replacements:  17500

----------------------------------------------------------------
Processing node 10: Align Retention Times
----------------------------------------------------------------
1. General Settings:
- Alignment Model:  Adaptive curve
- Alignment Fallback:  Use Linear Model
- Maximum Shift [min]:  0.1
- Shift Reference File:  True
- Mass Tolerance:  5 ppm
- Remove Outlier:  True

----------------------------------------------------------------
Processing node 9: Detect Unknown Compounds
----------------------------------------------------------------

1. General Settings:
- Mass Tolerance [ppm]:  5 ppm
- Intensity Tolerance [%]:  30
- S/N Threshold:  2
- Min. Peak Intensity:  100000
- Ions:
  [2M+ACN+H]+1
  [2M+ACN+Na]+1
  [2M+FA-H]-1
  [2M+H]+1
  [2M+K]+1
  [2M+Na]+1
  [2M+NH4]+1
  [2M-H]-1
  [2M-H+HAc]-1
  [M+2H]+2
  [M+3H]+3
  [M+ACN+2H]+2
  [M+ACN+H]+1
  [M+ACN+Na]+1
  [M+Cl]-1
  [M+DMSO+H]+1
  [M+FA-H]-1
  [M+H]+1
  [M+H+K]+2
  [M+H+MeOH]+1
  [M+H+Na]+2
  [M+H+NH4]+2
  [M+H-H2O]+1
  [M+H-NH3]+1
  [M+K]+1
  [M+Na]+1
  [M+NH4]+1
  [M-2H]-2
  [M-2H+K]-1
  [M-H]-1
  [M-H+HAc]-1
  [M-H+TFA]-1
  [M-H-H2O]-1
- Base Ions:  [M+H]+1; [M-H]-1
- Min. Element Counts:  C H
- Max. Element Counts:  C90 H190 Br3 Cl4 K2 N10 Na2 O15 P2 S5

2. Peak Detection:
- Filter Peaks:  True
- Max. Peak Width [min]:  0.5
- Remove Singlets:  True
- Min. # Scans per Peak:  7
- Min. # Isotopes:  2

-----------------------------------------------------------------
Processing node 8: Group Unknown Compounds
-----------------------------------------------------------------
1. Compound Consolidation:
- Mass Tolerance:  5 ppm
- RT Tolerance [min]:  0.1

2. Fragment Data Selection:
- Preferred Ions:  [M+H]+1; [M-H]-1

-----------------------------------------------------------------
Processing node 22: Search mzCloud
-----------------------------------------------------------------
1. Search Settings:
- Compound Classes:  All
- Match Ion Activation Type:  True
- Match Ion Activation Energy:  Any
- Ion Activation Energy Tolerance:  100
- Apply Intensity Threshold:  False
- Precursor Mass Tolerance:  5 ppm
- FT Fragment Mass Tolerance:  10 ppm
- IT Fragment Mass Tolerance:  0.1 Da
- Identity Search:  HighChem HighRes
- Similarity Search:  Similarity Forward
- Library:  Reference
- Post Processing:  Recalibrated
- Match Factor Threshold:  0
- Max. # Results:  20

-----------------------------------------------------------------
Processing node 16: Predict Compositions
-----------------------------------------------------------------
1. Prediction Settings:
- Mass Tolerance:  5 ppm
- Min. Element Counts:  C H
- Max. Element Counts:  C90 H190 Br3 Cl4 N10 O15 P2 S5
- Min. RDBE:  -1
- Max. RDBE:  40
- Min. H/C:  0.1
- Max. H/C:  3
- Max. # Candidates:  10
- Max. # Internal Candidates:  500

2. Pattern Matching:
- Intensity Tolerance [%]:  30
- Intensity Threshold [%]:  0.1
- S/N Threshold:  3
- Min. Spectral Fit [%]:  10
- Min. Pattern Cov. [%]:  90
- Use Dynamic Recalibration:  True

3. Fragments Matching:
- Use Fragments Matching:  True
- Mass Tolerance:  5 ppm
- S/N Threshold:  3

-----------------------------------------------------------------

Processing node 23: Search ChemSpider

-----------------------------------------------------------------

1. Search Settings:
- Mass Tolerance:  5 ppm
- Database(s):
    BioCyc
    Cayman Chemical
    ChEBI
    FDA UNII - NLM
    Human Metabolome Database
    KEGG
    LipidMAPS
    MCISB
    SMPDB Small Molecule Pathway Database
- Max. # of results per compound:  100
- Max. # of Predicted Compositions to be searched per Compound:  5
- Result Order (for Max. # of results per compound):  Order By Reference Count (DESC)

2. Predicted Composition Annotation:
- Check All Predicted Compositions:  False

-----------------------------------------------------------------

Processing node 27: Search Mass Lists

-----------------------------------------------------------------

1. Search Settings:
- Input file(s):  IEM_4400_AAAC_database_V2.0.csv
- Mass Tolerance:  5 ppm
- Show extra Fields as Columns:  True
- Consider Retention Time:  True
- RT Tolerance :  1

-----------------------------------------------------------------

Processing node 20: Mark Background Compounds

-----------------------------------------------------------------

1. General Settings:
- Max. Sample/Blank:  5
- Max. Blank/Sample:  0
- Hide Background:  True

-----------------------------------------------------------------

Processing node 28: Create Mass Trace

-----------------------------------------------------------------

1. General Settings:
- Trace Type:  TIC
- MS Order:  MS1
- Polarity:  +
- Custom Label:  (not specified)

2. XIC Settings:
- Mass [Da]:  0
- Mass Tolerance:  5 ppm

-----------------------------------------------------------------
Processing node 17: Differential Analysis
-----------------------------------------------------------------
1. General Settings:
- Log10 Transform Values:  True

-----------------------------------------------------------------
Processing node 24: Descriptive Statistics
-----------------------------------------------------------------
No parameters

-----------------------------------------------------------------
Processing node 29: Assign Compound Annotations
-----------------------------------------------------------------
1. General Settings:
- Mass Tolerance:  5 ppm

2. Data Sources:
- Data Source #1:  mzCloud Search
- Data Source #2:  (not specified)
- Data Source #3:  (not specified)
- Data Source #4:  (not specified)
