## Supplementary figures and images for "Studying autism using untargeted metabolomics in newborn screening samples"

### FigureS1_Analysis_Outliers

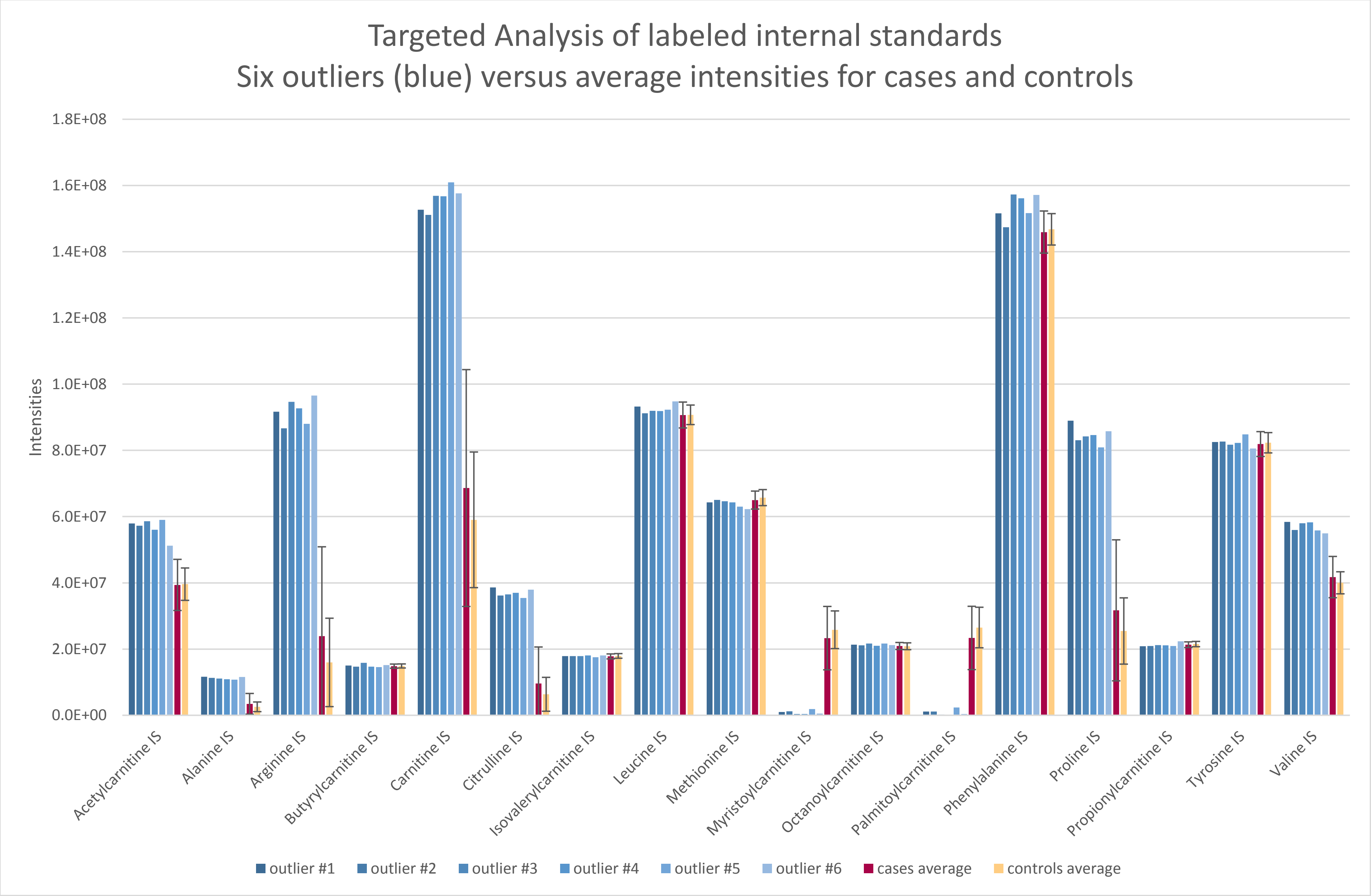

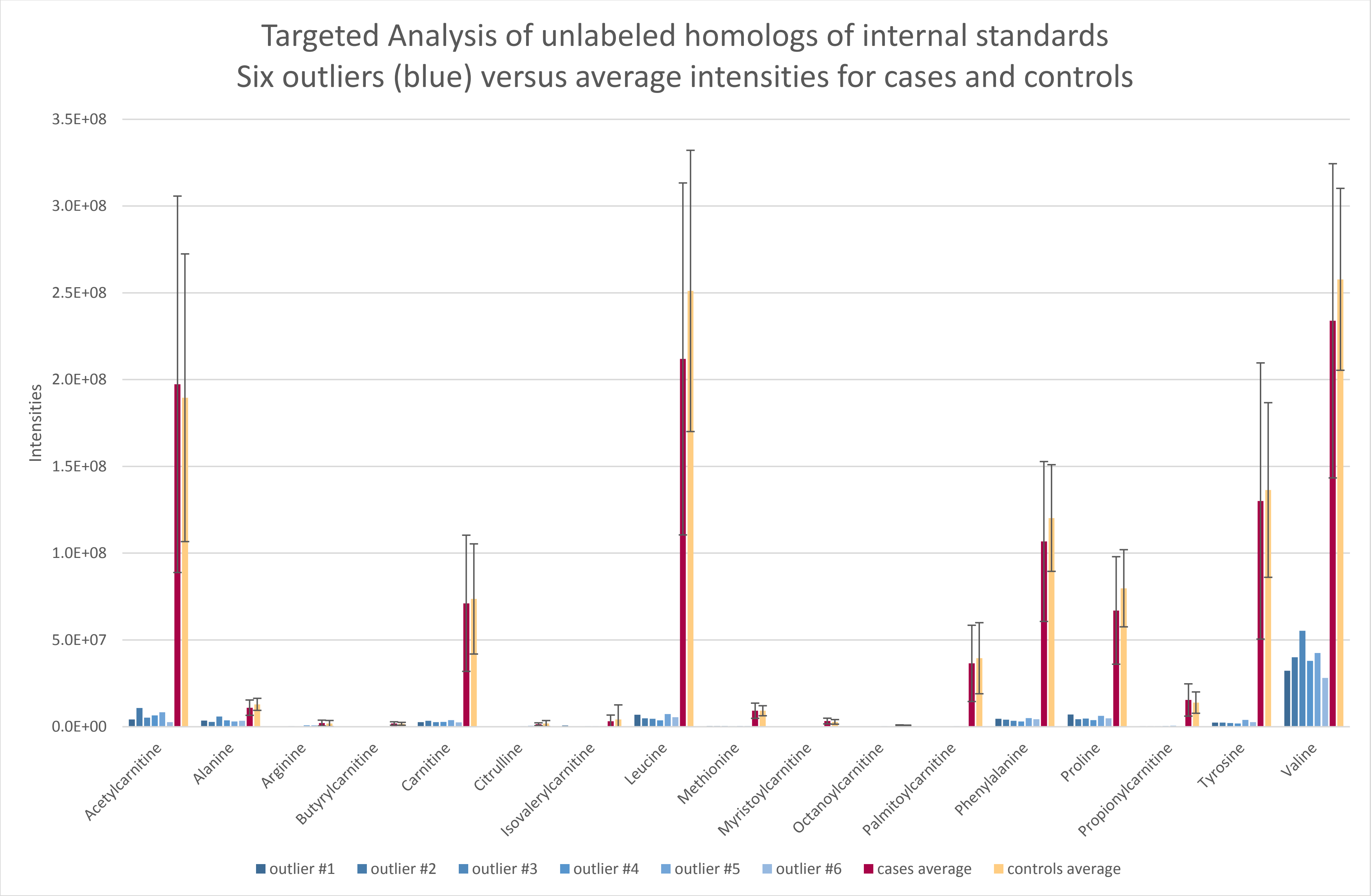

#### Heatmap of untargeted analysis features intensities generated using MetaboAnalyst 4.0

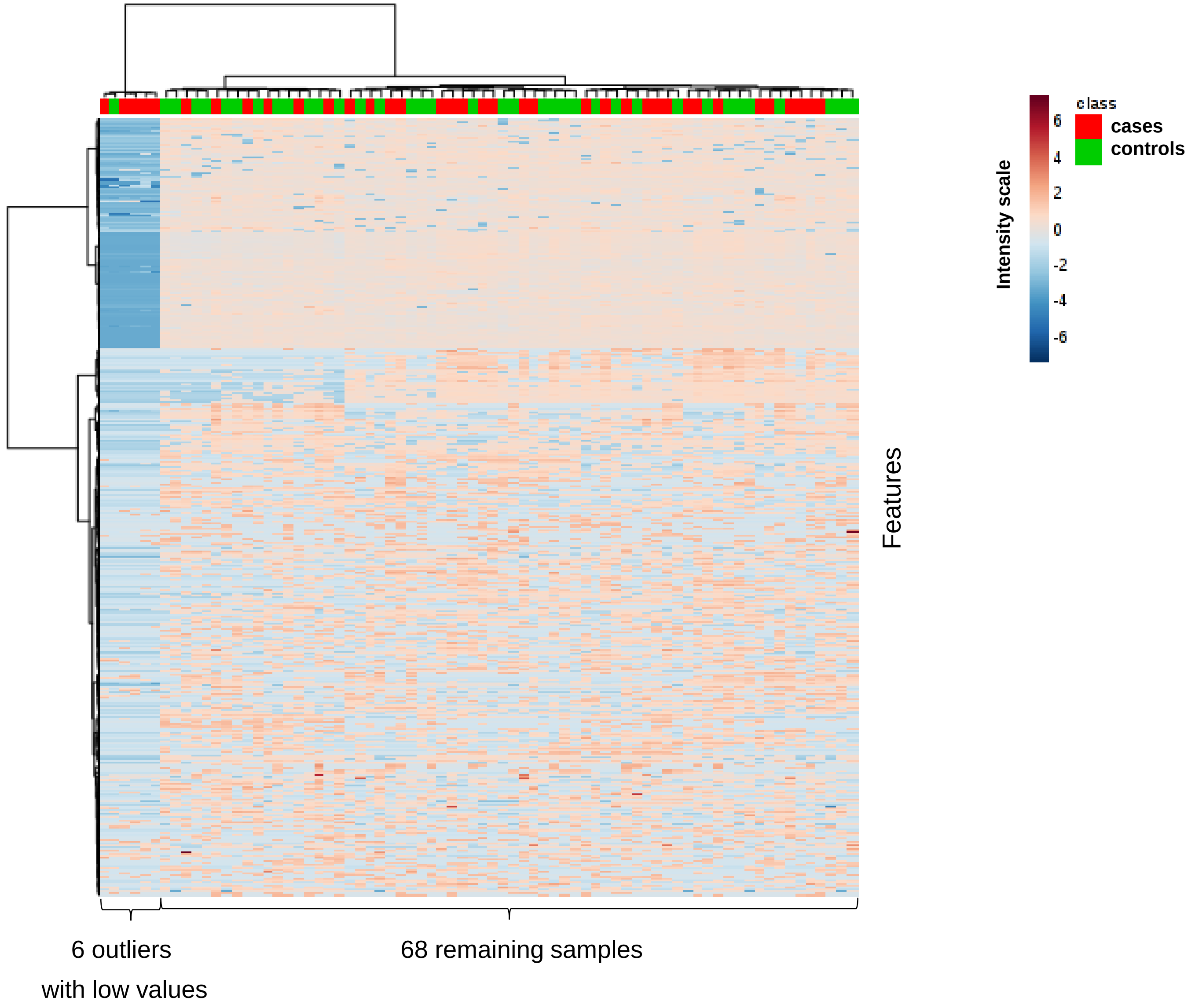
