## Supplementary material for "Studying autism using untargeted metabolomics in newborn screening samples": TableS5_Fragmentation_profiles

**Fragmentation profiles of the two unknown features to be monitored in future studies as well as methacholine as shown in Table 2.**

The fragment masses and intensities were extracted from the aggregated .mgf file computed by MZmine and used in the network analysis.

| ID8605 selected based on fold change | ID5593 selected based on significant p values before FDR correction | Methacholine selected based on significant p values before FDR correction |
|---|---|---|
| BEGIN IONS<br>FEATURE_ID=8605<br>PEPMASS=1014.4892<br>SCANS=8605<br>RTINSECONDS=398.449<br>CHARGE=1+<br>MSLEVEL=2<br>99.4840 5.0E3<br>130.2113 4.7E3<br>178.2334 6.2E3<br>498.1496 2.9E5<br>557.1633 3.9E5<br>616.1768 1.3E6<br>END IONS | BEGIN IONS<br>FEATURE_ID=5593<br>PEPMASS=231.1701<br>SCANS=5593<br>RTINSECONDS=166.747<br>CHARGE=1+<br>MSLEVEL=2<br>72.0808 4.2E4<br>84.9599 4.4E3<br>103.8799 2.1E3<br>144.7223 2.3E3<br>END IONS | BEGIN IONS<br>FEATURE_ID=159<br>PEPMASS=160.1332<br>SCANS=159<br>RTINSECONDS=26.97<br>CHARGE=1+<br>MSLEVEL=2<br>55.0546 2.1E5<br>59.0493 5.7E4<br>60.0810 6.1E5<br>101.0598 1.9E6<br>131.9749 2.3E4<br>END IONS |
